# Redefining Clinical Hyperprogression: the Incidence, Clinical Implications, and Risk Factors of Hyperprogression in Non-Small Cell Lung Cancer Treated with Immunotherapy

**DOI:** 10.1101/2024.01.04.23300116

**Authors:** Trie Arni Djunadi, Youjin Oh, Jeeyeon Lee, Jisang Yu, Liam Il-Young Chung, Yeunho Lee, Leeseul Kim, Timothy Hong, Soowon Lee, Zunairah Shah, Joo Hee Park, Sung Mi Yoon, Young Kwang Chae

**Affiliations:** Feinberg School of Medicine, Northwestern University, Chicago, IL, USA; School of Medicine, Kyungpook National University, Daegu, Republic of Korea; University of Hawai’i, Honolulu, Hawai’i, USA; Ascension Saint Francis Hospital, Evanston, IL, USA; Northwestern University, Evanston, IL, USA; Baylor University, St, Waco, TX, USA

**Keywords:** Respiratory tract tumours, Risk Factors, Retrospective Studies, Prognosis, Immunology

## Abstract

**Introduction:** Immune checkpoint inhibitors (ICIs) may be associated with hyperprogressive disease (HPD). However, there is currently no standardized definition of HPD, with its risk factors and clinical implications remaining unclear. We investigated HPD in lung cancer patients undergoing immunotherapy, aiming to redefine HPD, identify risk factors, and assess its impact on survival.

**Methods:** Clinical and radiologic data from 121 non-small cell lung cancer (NSCLC) patients with 136 immunotherapy cases were reviewed retrospectively. Three HPD definitions (Champiat et al., HPDc; Saâda-Bouzid et al., HPDs; and Ferrara et al., HPDf) were employed. Additionally, all new measurable lesions on the post-treatment CT scan were incorporated in measuring the sum of longest diameters (SLD) to define modified HPD (mHPD).

**Results:** Among the 121 patients, 4 (3.3%) had HPDc, 11 (9.1%) had HPDs, and none had HPDf. Adding all new measurable lesions increased HPD incidence by 5%–10% across definitions. Multivariate analysis revealed significantly lower progression-free survival (PFS) and overall survival (OS) for patients with HPDc (HR 5.25, p=0.001; HR 3.75, p=0.015) and HPDs (HR 3.74, p<0.001; HR 3.46, p<0.001) compared to those without. Patients with mHPD showed similarly poor survival outcomes as HPD patients. Liver metastasis at diagnosis was associated with HPDs, and a high tumor burden correlated with HPDc.

**Conclusions:** The incidence and risk factors of HPD varied with different definitions, but mHPD identified more cases with poor outcomes. This comprehensive approach may enhance the identification of at-risk patients and lead to a better understanding of HPD in lung cancer during immunotherapy.

## Introduction

Immune checkpoint inhibitors (ICI) have significantly improved the treatment of non-small cell lung cancer (NSCLC) by enhancing the anti-tumor immune response (1–3). Although immunotherapy is a major breakthrough in treating NSCLC, not all patients experience a favorable response. A potential adverse effect affecting approximately 5%–20% of NSCLC patients is hyperprogressive disease (HPD), characterized by an unusual increase in tumor burden following immunotherapy (4–7).

Various HPD definitions yield incidence ranging from 5% to 19.2%, reflecting differences in lesion inclusion criteria and tumor growth speed calculation (4, 5, 7–13). Some definitions consider target lesions only for a sum of longest diameters (SLD), while others include new lesions. Parameters for measuring tumor growth speed, such as tumor growth kinetics (TGK) and tumor growth rate (TGR), also vary (8, 10–14). For example, one study defines HPD as an over 50% increase in tumor burden with less than 2-month of time to treatment failure and more than a two-fold increase in progression pace (12), while other bases it on a two-fold TGR increase compared to pre-immunotherapy (8). Another research group defines HPD as disease progression with TGR increase exceeding 50% post-immunotherapy compared to pre-immunotherapy (11). These discrepancies lead to inconsistent findings across definitions.

The concept of HPD has been a topic of continued debate. Despite the efforts to define HPD as an independent clinical phenomenon associated with immunotherapy, some physicians regard HPD as a characteristic of cancer response or as part of rapid progression (13). Furthermore, there are limitations to the conventional definitions of HPD. When the definitions only consider target lesions, patients who experience accelerated tumor growth limited to new lesions may not be classified as having HPD (8, 10, 11, 15). This may lead to an unmet need in the proper identification and management of patients with HPD.

To better understand HPD in NSCLC, we investigated the incidence of HPD using various published definitions. Also, we redefined HPD by incorporating all new measurable lesions to measure the tumor burden and tumor growth more accurately. Survival outcomes and risk factors were also investigated for patients with HPD and mHPD.

## Materials and methods

This retrospective study was approved by the Institutional Review Board of Northwestern University (STU00207117). And the research methods were performed in accordance with the Declaration of Helsinki. It was not possible to involve patients or the public in the design, or conduct, or reporting, or dissemination plans of our research.

### Patient selection

We included patients with stage unresectable III-IV NSCLC who received immunotherapy at Northwestern Memorial Hospital between January 2014 and September 2021, either as monotherapy or combined with other treatments. Of the 261 patients treated during this period, patients who did not undergo a baseline or follow-up CT scan (n=67), had no target lesions that met the RECIST criteria (n=9), or underwent adjuvant immunotherapy after surgery (n=11) were excluded. Also, 53 patients were excluded as they did not have an eligible pre-baseline CT scan, which is necessary for calculating TGR or TGK. Finally, 121 patients with 136 immunotherapy cases were included for evaluation of HPD (Supplementary Figure 1).

### Clinicopathologic variables

Clinical variables evaluated include age at diagnosis, sex, smoking history, ECOG performance status (PS), tumor histology and stage at immunotherapy, programmed death (PD)-L1 status, tumor mutational burden (TMB), neutrophil-to-lymphocyte ratio (NLR), platelet count, line of immunotherapy treatment, and immunotherapy regimens.

Histologic types of NSCLC were divided into squamous and non-squamous cell carcinoma, and the non-squamous cell carcinoma was subdivided into adenocarcinoma, large cell carcinoma, poorly differentiated carcinoma, and NSCLC not otherwise specified (NOS). Although PD-L1 expression was defined as positive when it is ≥1% in immunohistochemical staining, the analysis was conducted with three groups: ≥50%, 1-49%, and <1%. Platelet, neutrophil, and lymphocyte counts were collected through complete blood counts and blood chemistry profiles, obtained immediately before immunotherapy. Platelet >450 (x10^9^/L), and the NLR >5 was considered high based on the previous literature (16–18). The line of immunotherapy was categorized as a first, second line, and third line or beyond, based on the sequence of immunotherapy within all the treatments administered to the patient. The immunotherapy regimen was divided into whether it contained chemotherapeutic agents or not. The number of metastatic lesions and occurrence of brain, bone, and liver metastasis at the time of initiating immunotherapy was also reviewed.

Genomic data was generated by next-generation sequencing (NGS) from 113 patients. NGS assay was performed in various laboratories, including Personal Genome Diagnostics (Labcorp, Burlington, NC), Altera (Natera, Austin, TX), and Tempus xT (Tempus, Chicago, IL). We compared the frequency of each alteration between patients with HPD and without HPD.

### Definitions of HPD

Among various definitions of HPD (19), we used three major definitions which were proposed by *Champiat* et al. (HPDc) (8), by *Saâda-Bouzid* et al. (HPDs) (10), and by *Ferrara* et al. (HPDf) (11) (Supplementary Table 1). HPDc was defined when the tumor has progressed based on RECIST evaluation and TGR-exp/TGR-ref (TGR ratio)>2; HPDs used only TGK-exp/TGK-ref (TGK ratio)>2; and HPDf used tumor progression based on RECIST and TGR-exp-TGR-ref>50%. However, one study that defined HPD based on immune-related response criteria (irRC) was excluded from this study because new lesions were already incorporated in the evaluation of tumor response to immunotherapy (12). Additionally, the irRC calculates the sum of the products of the two largest perpendicular diameters, whereas the RECIST measures the SLD (20). Three definitions of HPD included in the study were re-evaluated by adding all new measurable lesions which were detected on the first post-treatment CT scan, and we defined them as mHPD.

### Treatment response and survival outcomes

Serial CT scans were obtained at three time points and reviewed by four physicians independently: pre-baseline 2 weeks to 3 months before immunotherapy initiation, baseline within 2 weeks before and 1 month after starting immunotherapy, and post-baseline within 3 to 6 months of immunotherapy initiation. The number and size of each target lesion, non-target lesion, and newly appearing lesion at post-treatment CT were separately measured and classified based on RECIST 1.1 criteria and other parameters. Based on RECIST 1.1, tumor responses to immunotherapy were classified as complete response (CR), partial response (PR), stable disease (SD), and progressive disease (PD). Additionally, in cases where the tumor response satisfied the criteria of HPD, the response was distinguished as PD and HPD. One additional CT scan at least three months after the first post-treatment CT scan was also reviewed to evaluate pseudoprogression. Overall survival (OS) was determined as the period from the start date of immunotherapy to the date of death from any cause. Progression-free survival (PFS) was defined as the time between the initiation of immunotherapy and evidence of disease progression or death (whichever comes first). For PFS, progression was determined for each patient based on radiologic evidence for progression according to RECIST evaluation. Progression was also considered when evaluation of RECIST was not available or undetermined, and when the patient was thought to be experiencing clinical progression. The tumor burden was defined as SLD of target lesions according to RECIST 1.1 and the objective response rate was defined as the percentage of people who had a partial response or complete response during immunotherapy.

### Statistical analysis

To evaluate risk factors associated with HPD or mHPD, the categorical variables were analyzed using Fisher’s exact or Chi-squared test, and the continuous variables were analyzed using independent samples t-test. The Kaplan–Meier method was used for survival analysis to estimate PFS and OS, and the Cox proportional hazard regression was performed to determine the association between clinicopathological factors and PFS or OS. For OS, multivariate analysis was performed to analyze the prognostic factors for OS. In the multivariate analysis, age, sex, smoking history, and other prognostic factors having p value less than 0.1 in univariate analysis were included. The model with a better fit was selected according to the Akaike information criterion. Comparison between the two or more groups (i.e., those without HPD, those with HPDc or HPDs, and those with mHPDc or mHPDs) was conducted using the log-rank test. Statistical analyses were performed using R version 4.2.2.

## Results

### Clinicopathologic characteristics

The mean age of patients was 67.3 years (SD, ±11.1), and 102 patients (84.3%) had stage IV NSCLC. Adenocarcinoma was the most common histologic type (n=90, 74.3%), and PD-L1 was positive among 69.6% (n=48) of the tumors. The mean tumor burden at baseline, calculated by the SLD of the target lesions according to RECIST criteria, was 68.5 mm (SD, ±39.9). A total of 31 patients (25.6%) were found to have ≥3 metastatic lesions, with brain, bone, and liver metastasis observed in 32 (26.4%), 33 (27.3%), and 27 (22.3%) patients, respectively (Supplementary Table 2).

The median follow-up period was 61 weeks (interquartile range [IQR] 24–116 weeks), and patients underwent post-treatment CT after immunotherapy at a median of 9 weeks (IQR 6–13 weeks). The objective response rate at the first post-treatment CT was 7.4% (n=9), while 32 patients (26.4%) had PD.

### Incidence of HPD and mHPD based on three different definitions

The incidences of HPD varied depending on the definitions used [HPDc (n=4, 3.3%); HPDs (n=11, 9.1%); HPDf (n=0, 0.0%)]. After adding the new measurable lesions to assess the HPD, the incidence increased by 5%-10% [mHPDc (n=14, 11.6%); mHPDs (n=17, 14.0%); and mHPDf (n=12, 9.9%)]. Eleven patients (9.1%) were consistently diagnosed with mHPD across all three modified definitions (Supplementary Figure 2A). When the incidence of HPD and mHPD was assessed in 136 immunotherapy cases, mHPD increased by 6%-11% [mHPDc (n=18, 13.2%); mHPDs (n=22, 16.2%); and mHPDf (n=16, 11.8%)] (Supplementary Figures 2B and 2C). There were 19 patients who had HPD or mHPD, and we assessed the changes in tumor size of target lesions and new measurable lesions at each time point based on RECIST 1.1 criteria (Supplementary Table 3). Among these, seven patients were eligible for evaluation of potential pseudoprogression. The remaining cases were excluded because follow-up CT scans at least three months from the post-treatment CT scans were not available. All seven patients did not show pseudoprogression.

The inclusion of new measurable lesions in assessing HPD had a significant impact on the TGR and TGK ratios, as shown in Supplementary Figure 3. For HPDc and mHPDc, the median TGR ratios were 2.5 (range 2.0-7.7) and 9.3 (range 2.0-255.0), respectively, indicating an approximately four-fold increase in the TGR ratio for mHPDc. In particular, one case showed the TGR ratio change from 0.09 to 255. The TGK ratio also increased, though to a lesser extent, from 3.0 (range 2.1-8.0) for HPDs to 4.3 (range 2.7-29.0) for mHPDs.

Post-treatment CT scans revealed a median tumor burden of 159.5 mm (range 88.0-251.0) for HPDc and 124.0 mm (range 21.0-251.0) for HPDs. However, with the inclusion of new measurable lesions in tumor burden, the median tumor burden rose to 213.0 mm (range 171.0-274.0) for HPDc and 171.0 mm (range 21.0-309.0) for HPDs, as shown in Supplementary Figure 4. Under both definitions, this represented an approximate growth of 25% in tumor burden.

### Association between HPD and survival outcomes

HPD was associated with worse survival outcomes, including PFS and OS in patients receiving immunotherapy for NSCLC across the definitions. Three groups were analyzed: the ‘non-PD’ group (non-PD), ‘HPD’ group (HPD), and ‘PD without HPD’ group (non-HPD-PD). Both HPD and non-HPD-PD groups showed significantly worse PFS and OS than the non-PD group. The risk for progression was higher in HPDc and HPDs than non-HPDc-PD and non-HPDs-PD [HPDc: HR 11.47 (95%-CI 4.29-30.70) vs. non-HPDc-PD: HR 2.02 (95%-CI 1.31-3.13); HPDs: HR 5.00 (95%-CI 2.72-9.19) vs. non-HPDs-PD: HR 1.88 (95%-CI 1.18-3.00)] (Figures 1A and 1B). The same pattern was observed in OS among the three groups. HPDc and HPDs demonstrated a higher HR for OS compared to the groups with non-HPDc-PD and non-HPDs-PD, respectively, though there was no statistical significance (Figures 1C and 1D). When the patients were categorized into four groups: non-PD, HPD, mHPD, and non-HPD/mHPD-PD, both the HPD and mHPD groups demonstrated a worse prognosis compared to the non-PD group (Figure 2). However, there was no significant difference in PFS and OS between patients with HPD and those with mHPD (Supplementary Figure 5).

**Figure 1.**
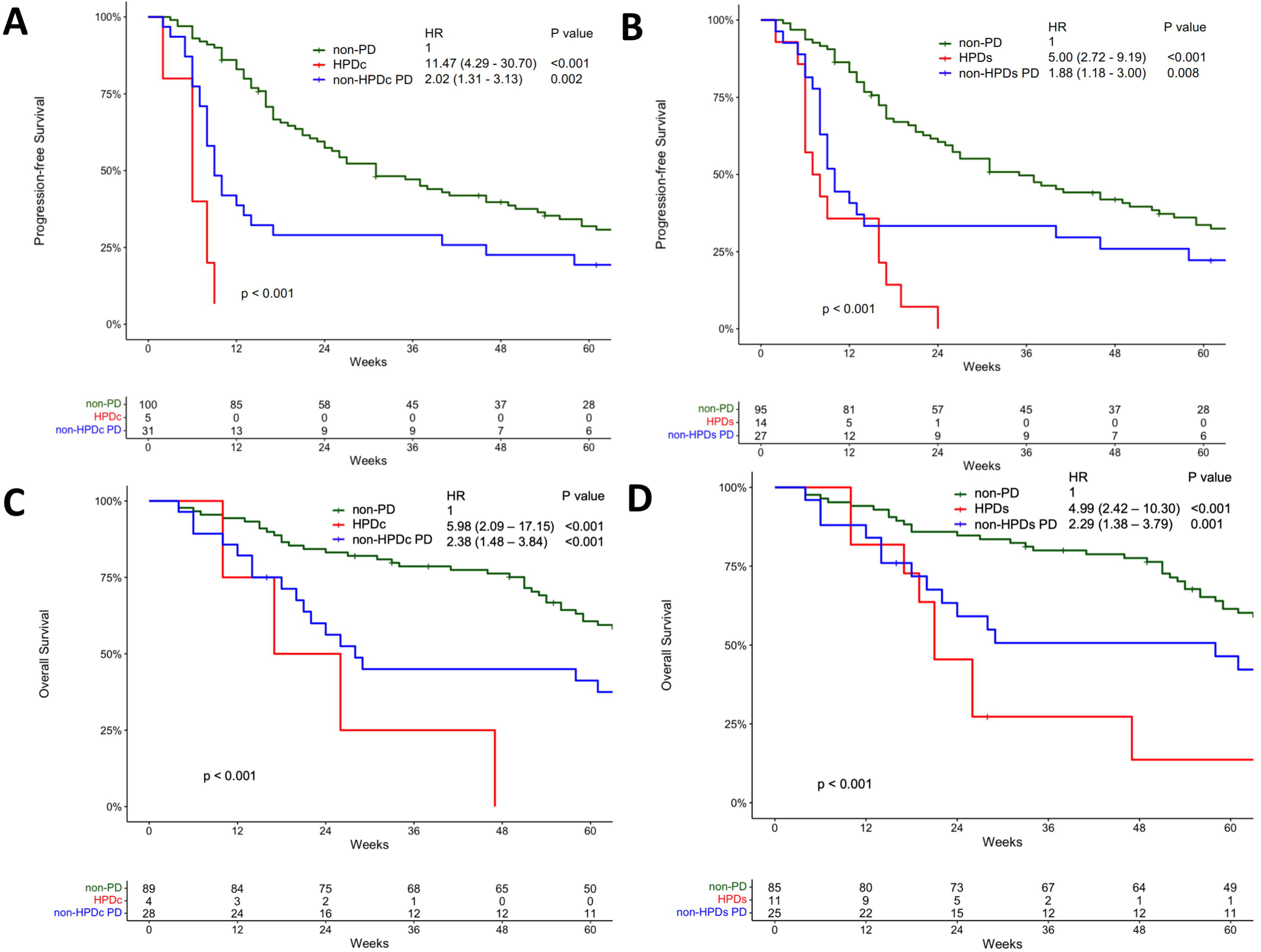
Progression-free survival (PFS) and overall survival (OS) among patients who received immunotherapy for non-small cell lung cancer. (A, C) PFS and OS in patients without progressive disease (non-PD), those with hyperprogression (HPDc), and those with PD but not HPD (non-HPDc PD) based on HPDc. (B, D) PFS and OS in patients without progressive disease (non-PD), those with hyperprogression (HPDs), and those with PD but not HPD (non-HPDs PD) based on HPDs. HPDc and HPDs, HPD according to the definition proposed by *Champiat* et al <u>[8]</u> and *Saâda-Bouzid* et al <u>[10]</u>.

**Figure 2.**
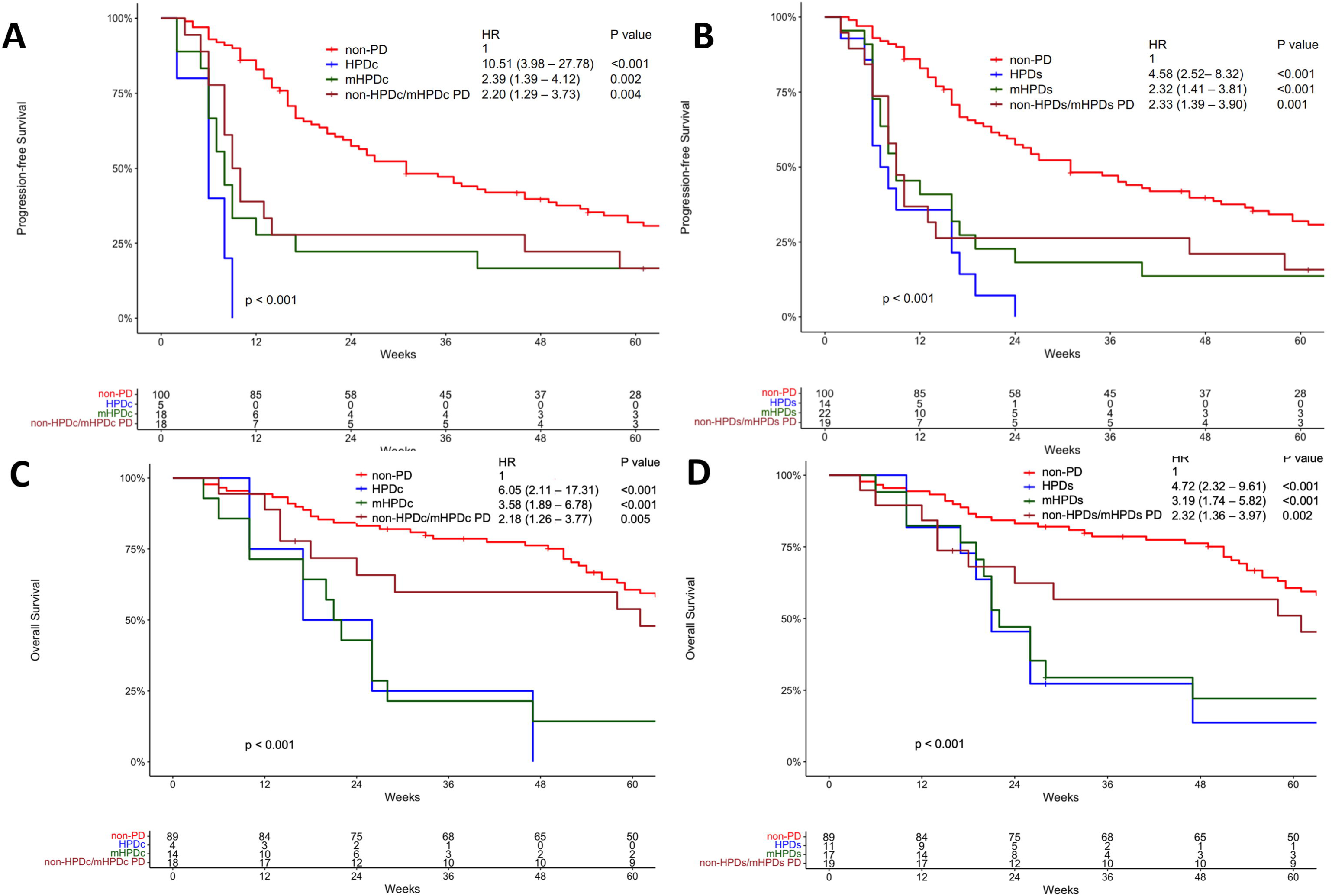
Progression-free survival (PFS) and overall survival (OS) among patients who received immunotherapy for non-small cell lung cancer. (A, C) PFS and OS in patients without progressive disease (non-PD), those with hyperprogression based on original and modified definition (HPDc and mHPDc), and those with PD but not HPD (non-HPDc PD) based on HPDc. (B, D) PFS and OS in patients without progressive disease (non-PD), with those with hyperprogression based on original and modified definition (HPDs and mHPDs), and those with PD but not HPD (non-HPDs PD) based on HPDs. HPDc and HPDs, HPD according to the definition proposed by *Champiat* et al <u>[8]</u> and *Saâda-Bouzid* et al <u>[10]</u>.

For PFS with 136 cases based on immunotherapy regimens, the following factors were observed to be associated with increased risk for progression: female (p=0.019), smoking history (p=0.059), presence of liver and bone metastasis (p=0.002 and p=0.027), HPDc (p<0.001) and HPDs (p<0.001). For OS with 121 patients, statistical significance was found in female patients (p=0.055), HPDc (p=0.005), and HPDs (p<0.001). In multivariate analysis, HPDc (p=0.015) and HPDs (p<0.001) were significantly associated with shorter OS (Table 1).

**Table 1.**
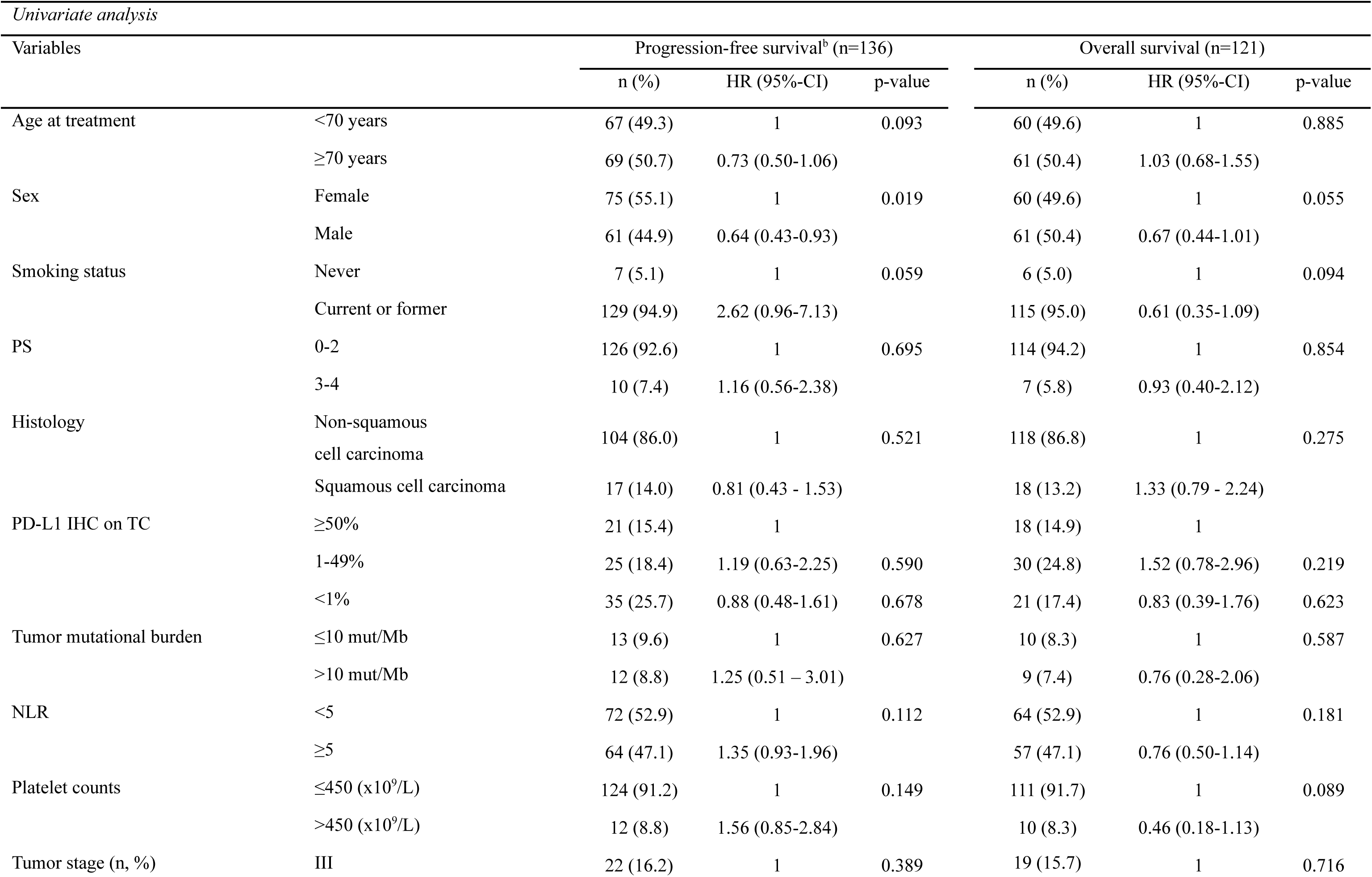
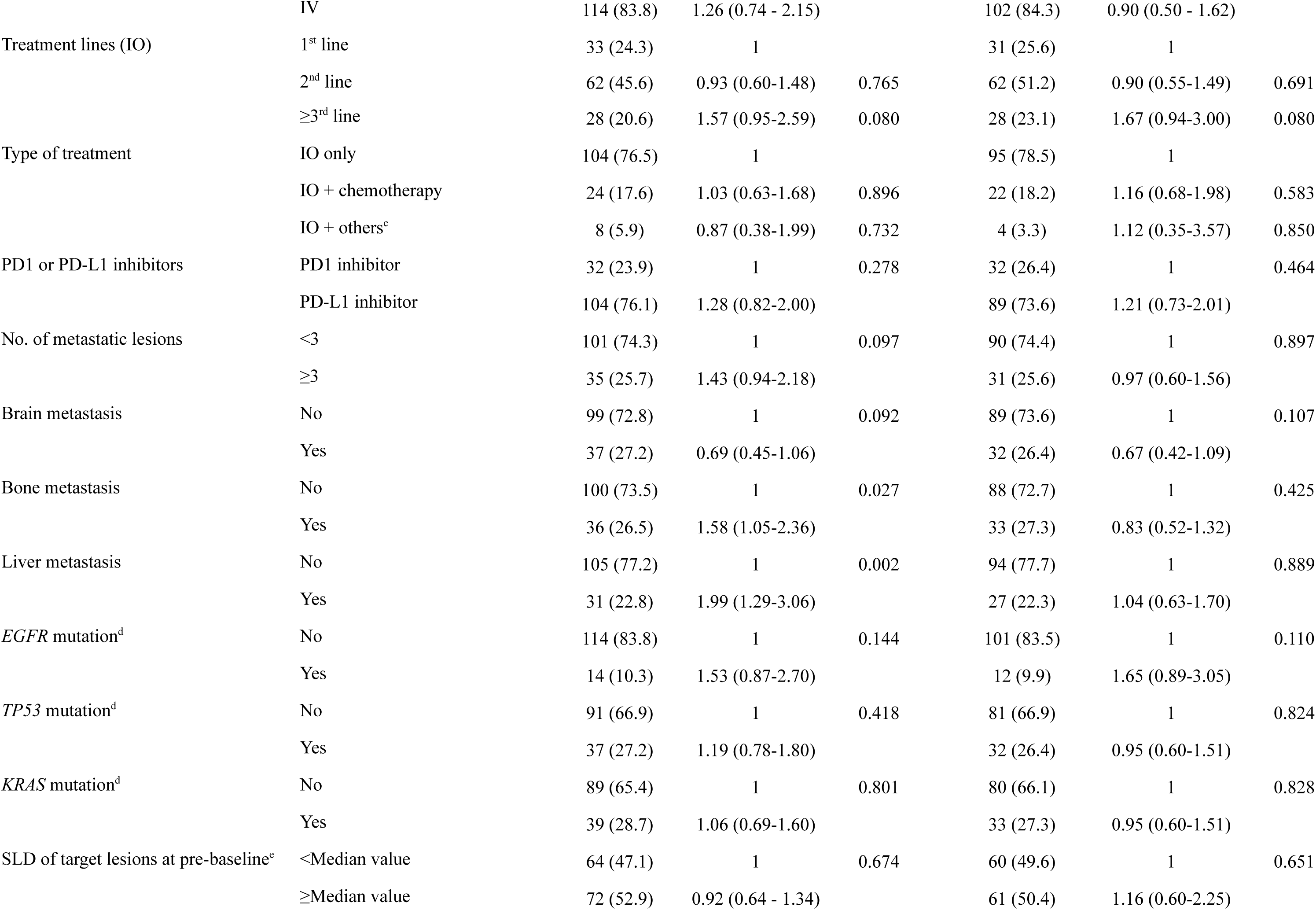
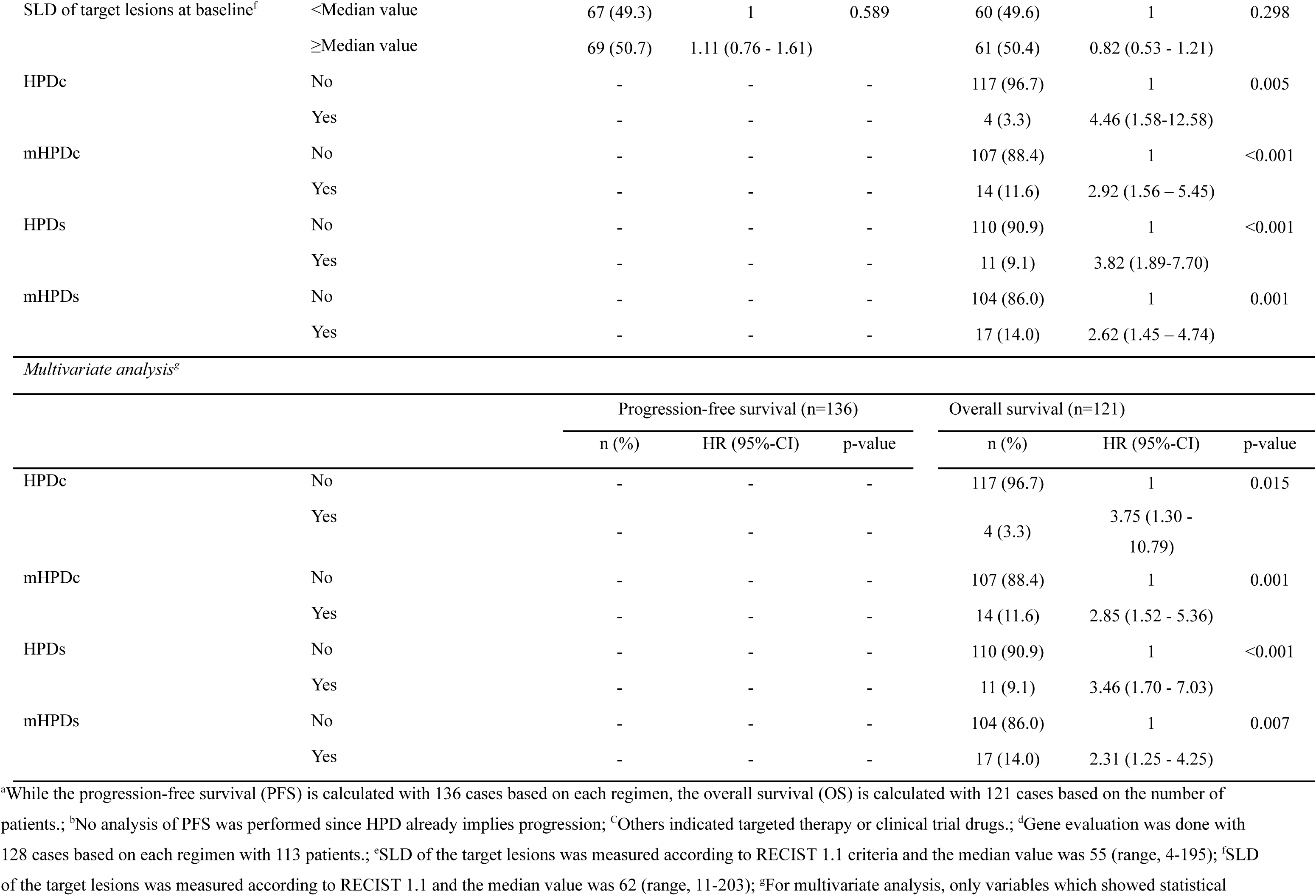
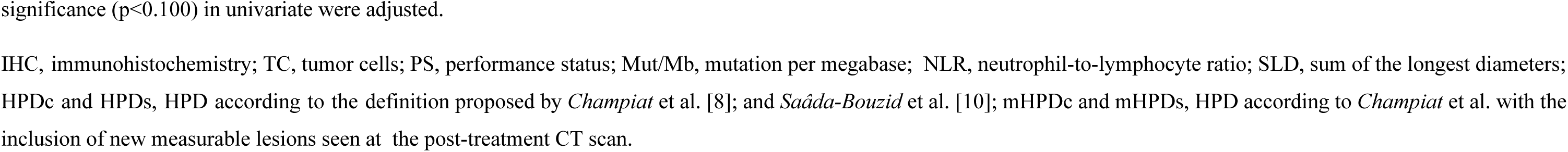
Univariate and multivariate analysis result for progression-free survival and overall survival in patients who received immunotherapy for non-small cell lung cancer

### Risk factors for HPD

There was no common risk factor between HPDc and HPDs. The SLD of target lesions at pre-baseline and baseline were significantly associated with HPDc (p=0.006 and 0.003), and the presence of liver metastasis at diagnosis was associated with HPDs (p=0.021) (Supplementary Table 2). However, for mHPD and non-mHPD, no significant risk factor was identified (Supplementary Table 4).

NGS results were available for only 113 patients in our study. Out of these, four had HPDc, and eight had HPDs. Fifty-two genomic alterations were found among these patients, comprising 49 somatic mutations and three copy number variations, as detailed in Supplementary Table 5. While the patients with HPDc had somatic mutations in the following seven genes (*KRAS, TP53, SMARCA4, STK11, NOTCH2, BRCA2*, and *BRAD1*) and amplifications in the following three genes (*AKT2, AXL*, and *MCL1*), the patients with HPDs had somatic mutations in additional five genes (*NOTCH1, FANCA, RAD50, FANCI,* and *GNAS*). Although the sample size was small, the single nucleotide variations of *BRAD1* and NOTCH2 and amplification in *AKT2, AXL*, and *MCL1* were associated with HPDc (p=0.010). However, no specific gene mutation or amplification was associated with HPDs (Supplementary Table 5).

### Prognostic implications of new lesions

Thirty-two patients (26.4%) demonstrated PD at the first post-treatment evaluation. After undergoing immunotherapy, 25 patients (78.1%) were confirmed to have developed additional new measurable lesions, and nine patients (28.1%) exhibited the development of ≥3 new lesions. OS for patients who developed ≥3 new measurable lesions following immunotherapy was significantly inferior to that of the group who had no new lesions or <3 new measurable lesions (p=0.046) (Figure 3).

**Figure 3.**
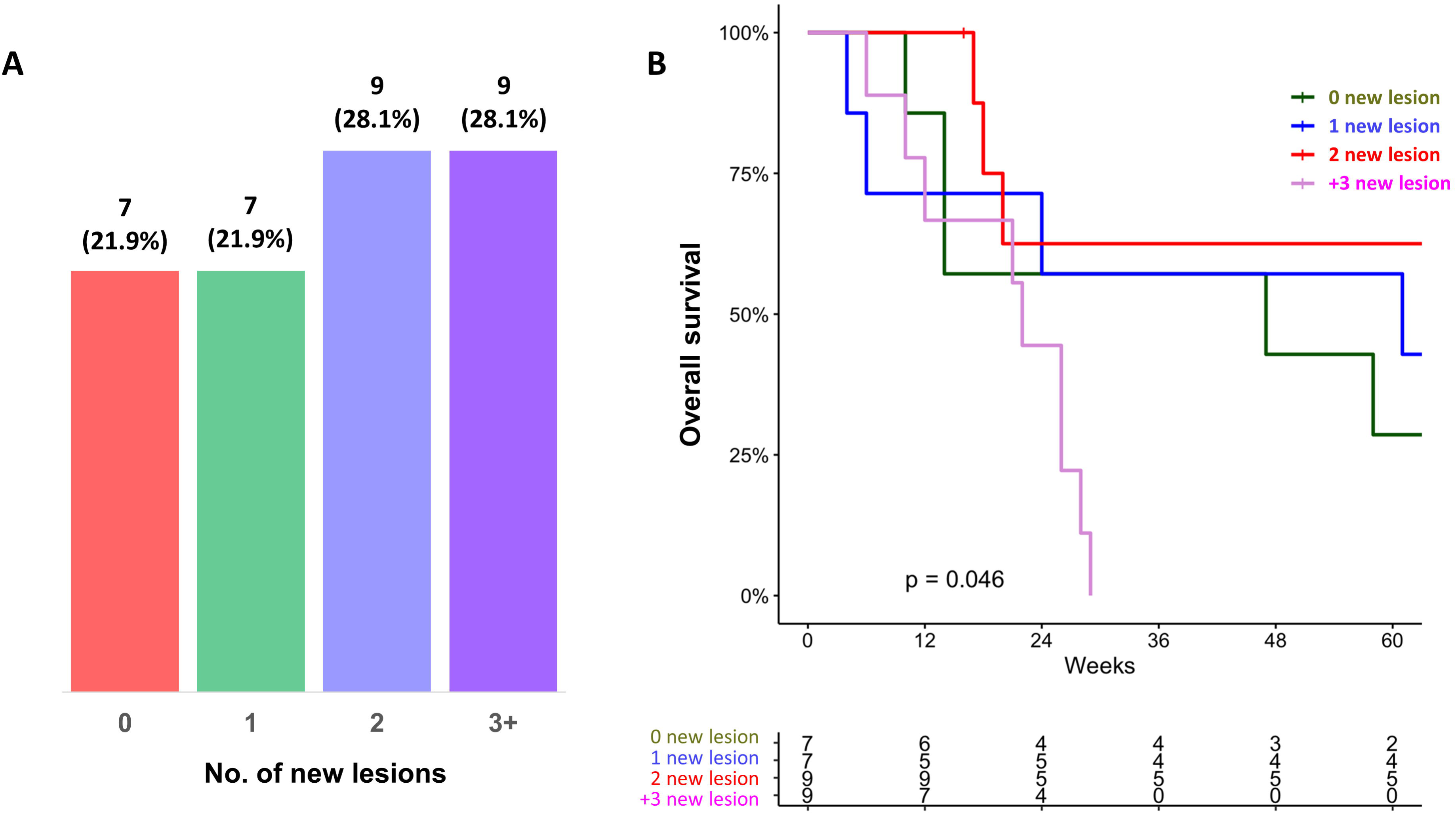
The proportion of new lesions and overall survival among the 32 patients who had progressive disease after immunotherapy at the first post-treatment evaluation. (A) The proportion of new lesions on the first CT after immunotherapy. (B) Comparison of overall survival according to the number of new lesions in patients with progressive disease.

Both PFS and OS were significantly worse in patients diagnosed with HPDc or HPDs, whether there were three or more new lesions (Figure 4). Compared to non-mHPD group, a significantly worse OS was only seen in those with ≥3 new lesions [mPHDc (≥3), HR 5.94, p<0.001; mHPDs (≥3), HR 6.09, p<0.001] (Figures 4G and 4H).

**Figure 4.**
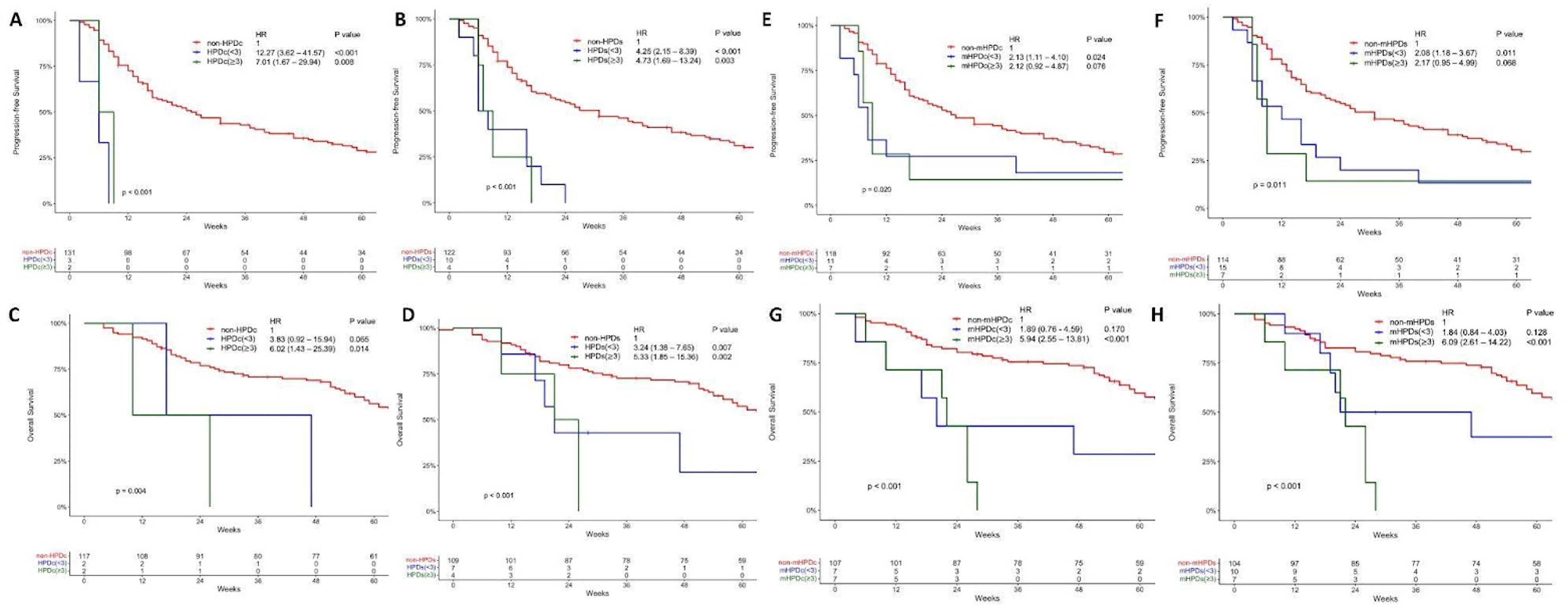
Progression-free survival (PFS) and overall survival (OS) in patients with or without hyperprogressive disease receiving immunotherapy for non-small cell lung cancer. (A, C) PFS and OS in patients with HPDc with <3 or ≥3 new lesions, and without HPDc. (B, D) PFS and OS in patients with HPDs with <3 or ≥3 new lesions, and without HPDs. (E, G) PFS and OS in patients with mHPDc with <3 or ≥3 new lesions, and without mHPDc. (F, H) PFS and OS in patients with mHPDs with <3 or ≥3 new lesions, and without mHPDs. HPDc and HPDs, HPD according to the definition proposed by *Champiat* et al <u>[8]</u> and *Saâda-Bouzid* et al <u>[10]</u>.

## Discussion

Our study revealed that the mHPD, which incorporates newly appearing measurable lesions on the first post-treatment CT, increased the incidence of HPD by 5%-10% across the different definitions. However, the survival outcomes of patients with mHPD were similarly poor compared to those with HPD defined by the original definition. Furthermore, patients with mHPD with ≥3 new measurable lesions had the worst OS among all cases. Our findings suggest incorporating new measurable lesions in treatment response assessment may help better assess HPD.

A higher HPD incidence was observed for the TGK ratio-based ‘HPDs’ than for the TGR ratio-based ‘HPDc’. Incidences of HPD in NSCLC patients treated with ICI monotherapy have varied, increasing in the order of ΔTGR, TGR ratio, and TGK ratio as follows 5.1% (n=12), 15.6% (n=37), and 18.1% (n=43) in one study (14), and 5.7% (n=8), 11.3% (n=16), and 17.0% (n=24) in the other study (7). However, two studies reported a similar proportion of NSCLC patients with HPD based on TGR ratio and TGK ratio (TGR ratio vs. TGK ratio: 20.5% (n=54) vs 20.4% (n=55), and 16.7% (n=7) vs. 16.7% (n=7), respectively) (21, 22). Notably, the definitions of HPD varied across these studies. In the two studies, TGK ratio-based HPD was defined among patients experiencing progression by RECIST 1.1, whereas, in our study, ‘HPDs’ was defined by TGK ratio regardless of RECIST 1.1 evaluation (14, 21). In other studies, TGR ratio-based HPD was defined regardless of RECIST 1.1 status, while in our study, TGR ratio-based ‘HPDc’ was defined in patients with progression by RECIST 1.1 (7, 22). Therefore, the prevalence of each definition after immunotherapy in NSCLC patients should be further investigated.

The incidence of HPD in our study was lower than in previous studies, which could be attributed to the high proportion of patients who received immunotherapy combined with other treatments, including chemotherapy (n=22, 18.2%), targeted therapy (n=2, 1.7%) and clinical trial drugs (n=2, 1.7%). Previous studies mainly included patients who received immunotherapy alone, with higher incidences ranging from 10% to 20% (7, 14, 21, 22). It has been reported that HPD occurs more frequently in patients receiving monotherapy with PD-1 and PD-L1 inhibitors compared to those treated with immunotherapy combined with chemotherapy in NSCLC (17.6% vs. 2.9%, p=0.031) (23). Furthermore, it has been suggested that combining chemotherapy and immunotherapy may be effective in preventing HPD as chemotherapy may synergize with immunotherapy to enhance the anti-tumor effect (24, 25).

Two factors were found to be associated with HPD. First, liver metastasis was an independent risk factor for ‘HPDs’. Previous studies have also highlighted liver metastasis as a risk factor for various HPD definitions in NSCLC consistently, such as TGR ratio ≥2 (7), ‘HPDc’ (8), and TGR ratio ≥2 and TGK ratio ≥2 with PD by RECIST 1.1 (21). In a recent meta-analysis, it was further demonstrated that NSCLC patients with liver metastasis were significantly more likely to develop HPD (HR 3.17, p<0.001), emphasizing its robustness as a risk factor (26). In the biopsy samples of melanoma patients treated with pembrolizumab, reduced CD8+ T cell in tumor margin was observed in patients with liver metastases compared with those without liver metastases, suggesting liver-induced immune tolerance (27). This may explain the strong correlation between liver metastases and HPD.

Secondly, a high tumor burden, measured by the SLD of target lesions according to RECIST 1.1 at pre-baseline/baseline, was identified as a risk factor for HPDc but not for ‘HPDs’ in our study. In the previous study, metabolic tumor burden, expressed by metabolic tumor volume and total lesion glycolysis measured by PET/CT, was associated with HPD in NSCLC patients (28). Tumor burden as primary lesion size was also significantly higher in HPD than in non-HPD patients with NSCLC (29). However, tumor burden measured by the sum of the RECIST 1.1 at baseline, as in our study, was not associated with HPD (8, 30). Further investigation is warranted given the heterogeneous results based on the different parameters for tumor burden.

‘HPDc’ and ‘HPDs’ were associated with worse OS than the non-HPD group. This finding aligns with previous studies that have consistently demonstrated a higher risk of death among patients with HPD in NSCLC: ‘HPDf’ (HR 2.18, p=0.003) (11), TGR ratio ≥2 and TGK ratio ≥2 (HR 5.079, p<0.001) (21), TTF ≤8 weeks (HR 2.48, p<0.001) (7), and TGK ratio ≥2 with PD and TTF <9 weeks (HR 2.66, p=0.009) (31). In our study, the association remained significant even after adjusting for clinical variables, including sex, and smoking history, regardless of the definition of HPD. Previous studies also reported a consistent association between HPD and worse OS in univariate and multivariate analyses (32).

Although various approaches and definitions have been proposed to evaluate HPD and tumor behavior, no consensus on the definition of HPD has yet to be reached (8, 10–12, 19). Current HPD definitions are based on RECIST criteria and do not consider the new lesions as relevant parameters for defining HPD. However, it has been suggested that newly appearing lesions after immunotherapy should also be considered, given that new lesions occur in more than 50% of patients with NSCLC regardless of treatment types during treatment (14, 33–35). Therefore, we investigated whether including new measurable lesions to assess HPD better can provide more clinical information and improve the prediction of survival outcomes.

Considering new lesions in defining HPD may allow for a more comprehensive assessment of the tumor burden in patients undergoing immunotherapy. An essential limitation of not considering new lesions is that patients who experience rapid tumor growth, primarily in newly developed lesions might not be classified as HPD (15). In our study, the inclusion of new lesions led to the diagnosis of HPD in an additional 10 (8.3%), 6 (5.0%), and 12 (10.0%) patients based on TGR ratio, TGK ratio, and ΔTGR, respectively. Furthermore, we observed that patients with mHPD had similarly poor survival outcomes as patients with HPD, and these associations persisted even after adjusting for confounding factors.

Additionally, having ≥3 new lesions was associated with worse overall survival outcomes than having two or fewer lesions. In a previous study, when new lesions were included in the definition of HPD, OS was significantly shorter among HPD patients than non-HPD patients (14). However, no significant difference in OS was observed when new lesions were not considered. Notably, the optimal threshold for the number of new lesions in predicting worse survival was two (14). These findings suggest that including new measurable lesions in the evaluation of HPD may provide value by identifying additional HPD patients who might require more attention.

It is equally important to distinguish pseudoprogression from HPD. On-treatment CT scans of the patients with HPD or mHPD were reviewed, and no pseudoprogression was detected. However, as pseudoprogression was reported in 0.6-5.8% of patients with advanced NSCLC (36), Distinguishing the two is of clinical significance. Therefore, following up on subsequent imaging studies and clinical evaluation are necessary (20, 37). When HPD is suspected in patients who show clinical improvement, pseudoprogression should be considered, and continuation of therapy for potential clinical benefit may be considered (38).

There are several proposed mechanisms underlying HPD. First, tumor-associated macrophages can boost tumor growth by binding to the Fc domain of PD-1 targeted monoclonal antibodies and modulating its functional activity (39). In an athymic mice model injected with human lung cancer cells, anti-PD-1 treatment accelerated tumor growth compared to the control group. However, anti-PD-1 antibodies lacking the Fc portion did not induce this tumor growth (39). Second, activated CD8+ T cells can trigger tumor growth by secreting interferon-gamma (INFγ), which leads to tumor growth by activating oncogenic pathways in tumor cells. HPD has been associated with increased expression of INFγ, fibroblast growth factor 2 (FGF2), MYC, and CD133 in melanoma and NSCLC patients undergoing immunotherapy, indicating the upregulated oncogenic stemness/invasiveness pathway (40). In a murine model depleting CD8+ T cells, HPD did not occur with anti-PD-L1 therapy (40). Third, PD-1 inhibitors can increase tumor-infiltrating immunosuppressive Treg cells expressing Ki67+ (eTreg). In gastric cancer patients treated with anti-PD-1 inhibitors, those with HPD exhibited an increased eTreg cells/CD8+ T cell ratio, whereas those without HPD showed a decreased ratio (41). Other mechanisms based on innate immune cells, including NK and dendritic cells, have also been reported (42–44).

This study possesses several notable strengths that contribute to its overall significance. First, it is the first study that applied all previously published definitions to redefine and comprehensively analyze patients with HPD. Second, it demonstrates the prognostic value of a modified definition incorporating the number of new lesions across different HPD definitions, which has not been previously explored. Third, while previous studies have identified the prognostic significance of the number of new lesions, our research further validates this finding specifically in the context of HPD. Fourth, we have effectively confirmed that the observed progression in our study is not attributed to pseudoprogression. However, it is important to acknowledge the limitations inherent in retrospective studies, also present in our study. Additionally, the relatively small sample size poses a constraint, which may introduce random errors and impact the generalizability of our findings.

In conclusion, our study shows that the incidence and risk factors of HPD differ depending on the definition used. The Incorporation of new measurable lesions into HPD definitions leads to the detection of more patients with HPD and consistently worse survival outcomes. Therefore, adding new measurable lesions in determining HPD may provide a more comprehensive definition that could more accurately reflect the extent of the tumor burden in patients undergoing immunotherapy. Our findings could help identify more patients who require careful monitoring during immunotherapy for NSCLC and potentially improve the survival outcomes of these patients.

## Data Availability

All data produced in the present work are contained in the manuscript

## List of Abbreviations

HPD: hyperprogressive disease
ICI: immune checkpoint inhibitors
NSCLC: non-small cell lung cancer
SLD: sum of longest diameters
TGK: tumor growth kinetics
CT: Computed tomography
TGR: tumor growth rate
HPDc: definitions suggested by *Champiat* et al.
HPDs: definitions suggested by *Saâda-Bouzid* et al.
HPDf: definitions suggested by *Ferrara* et al.
PFS: progression-free survival
OS: overall survival
mHPD: modified HPD
PS: ECOG performance status
PD-L1: programmed death L1
TMB: tumor mutational burden
NLR: neutrophil-to-lymphocyte ratio
NOS: not otherwise specified
irRC: immune-related response criteria
CR: complete response
PR: partial response
SD: stable disease
PD: progressive disease
CI: confidence interval
NGS: next-generation sequencing
INFγ: interferon-gamma
FGF2: fibroblast growth factor 2

## Declarations

### Ethics approval and consent to participate

This study was approved by the Institutional Review Board Committee of Northwestern University (STU00207117).

### Consent for publication

Consent was waived because only de-identified information was used

### Availability of data and material

The data sets generated and/or analyzed during the current study are not publicly available. However, they are available from the corresponding author upon reasonable request.

### Authors’ contributions

Conception and design: Young Kwang Chae.

Administrative support: Trie Arni Djunadi, Youjin Oh, Liam Il-young Chung. Provision of study materials or patients: Trie Arni Djunadi, Youjin Oh, Jeeyeon Lee.

Collection and assembly of data: Trie Arni Djunadi, Youjin Oh, Jeeyeon Lee, Jisang Yu, Joohee Park, Sungmi Yoon, Zunairah Shah, Soowon Lee, Timothy Hong.

Data analysis and interpretation: Young Kwang Chae, Youjin Oh, Trie Arni Djunadi, Jeeyeon Lee. Manuscript writing: All authors.

Final approval of manuscript: All authors. Accountable for all aspects of the work: All authors.

## Acknowledgments

Not applicable

## Supplementary figures legend

Supplementary Figure 1. Flowchart of the patients selection process.

Supplementary Figure 2. Incidence of hyperprogressive disease (HPD) according to the original and modified definitions. (A) Incidences in 121 patients who received immunotherapy for non-small cell lung cancer. (B) Incidences in 136 cases in which immunotherapy was given for non-small cell lung cancer. (C) Changes in the incidence of HPD and modified HPD (mHPD) according to the three definitions of HPD. HPDc, HPDs, and HPDf, HPD according to the definition proposed by *Champiat* et al <u>[8]</u>, *Saâda-Bouzid* et al <u>[10],</u> and *Ferrara* et al <u>[11]</u>.

Supplementary Figure 3. Changes of tumor growth rate (TGR) ratio (A) and tumor growth kinetics (TGK) ratio (B) using original definitions (HPDc, HPDs) and modified definitions (mHPDc, mHPDs). HPDc, HPDs, and HPDf, HPD according to the definition proposed by *Champiat* et al <u>[8]</u>, *Saâda-Bouzid* et al <u>[10],</u> and *Ferrara* et al <u>[11]</u>.

Supplementary Figure 4. Changes of tumor burden, measured as the SLD of the target lesions for using original definitions (A, C) and modified definitions (B, D). For the original definition, the SLD of the target lesions was measured according to RECIST 1.1. For modified definition, all new measurable lesions on the post-treatment CT scan were incorporated in measuring the sum of longest diameters (SLD). Each number represents each patient. (A, B) Tumor burden at each time point (pre-baseline, baseline, and post-baseline) by using HPDc and mHPDc. (C, D) Tumor burden at each time point using HPDs and mHPDs. HPDc, HPDs, and HPDf, HPD according to the definition proposed by *Champiat* et al [8], *Saâda-Bouzid* et al [10], and *Ferrara* et al <u>[11]</u>.

Supplementary Figure 5. Comparison of progression-free survival (PFS) and overall survival (OS) in patients with hyperprogressive disease (HPD) and modified HPD (mHPD) after immunotherapy for non-small cell lung cancer. (A, C) PFS and OS were compared between patients with HPDc and mHPDc. (B, D) PFS and OS were compared between patients with HPDs and mHPDs. HPDc and HPDs, HPD according to the definition proposed by *Champiat* et al <u>[8]</u> and *Saâda-Bouzid* et al <u>[10]</u>.

**Supplementary Table 1.**
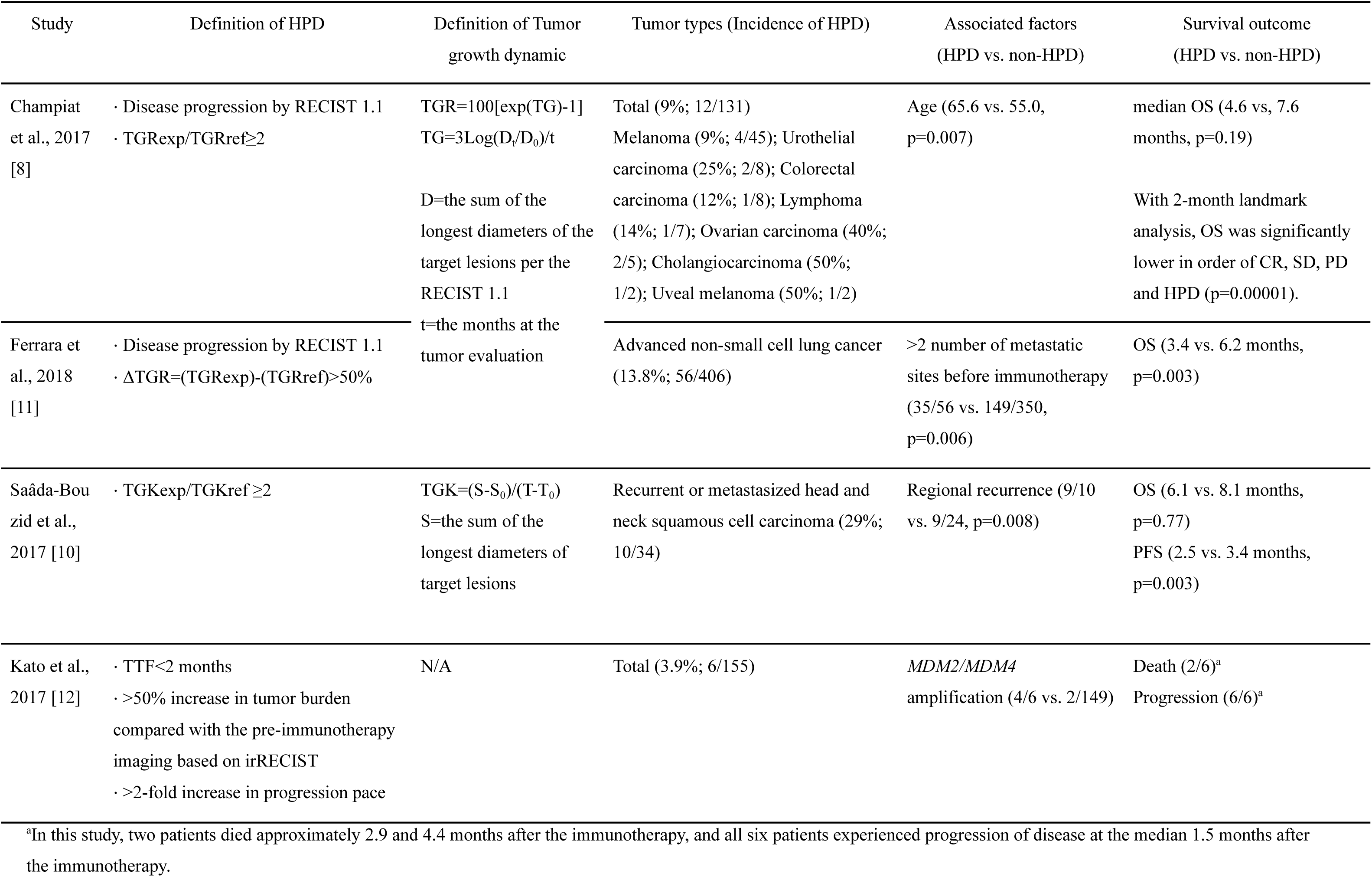
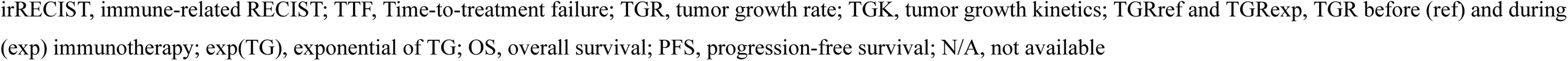
Summary of major definitions, associated factors, and survival outcome of hyperprogressive disease

**Supplementary Table 2.** Clinical and pathological characteristics between patients with non-small cell lung cancer (NSCLC) who were diagnosed with and without hyperprogressive disease (HPD) based on each definition

| Variables |  | Total<br>(n=121) | HPDc<br>(n=4) | non-HPDc<br>(n=117) | p-value | HPDs<br>(n=11) | non-HPDs<br>(n=110) | p-value |
| --- | --- | --- | --- | --- | --- | --- | --- | --- |
| Age (years, mean±SD) |  | 67.3±11.1 | 71.5±4.8 | 68.4±10.9 | 0.574 | 66.6±10.3 | 68.7±10.8 | 0.546 |
| Sex (n, %) | Female | 65 (53.7) | 3 (75.0) | 62 (53.0) | 0.720 | 8 (72.7) | 57 (51.8) | 0.313 |
|  | Male | 56 (46.3) | 1 (25.0) | 55 (47.0) |  | 3 (27.3) | 53 (48.2) |  |
| Smoking status (n, %) | Current | 6 (5.0) | 0 (0.0) | 6 (5.1) | 0.536 | 0 (0.0) | 6 (5.5) | 0.727 |
|  | Never | 22 (18.2) | 0 (0.0) | 22 (18.8) |  | 2 (18.2) | 20 (18.2) |  |
|  | Former | 93 (76.9) | 4 (100.0) | 89 (76.1) |  | 9 (81.8) | 84 (76.4) |  |
| PS (n, %) | 0-2 | 114 (94.2) | 4 (100.0) | 110 (94.0) | 1.000 | 11 (100.0) | 103 (93.6) | 0.853 |
|  | 3-4 | 7 (5.8) | 0 (0.0) | 7 (6.0) |  | 0 (0.0) | 7 (6.4) |  |
| Tumor stage (n, %) | III | 19 (15.7) | 0 (0.0) | 19 (16.2) | 0.858 | 0 (0.0) | 19 (17.3) | 0.286 |
|  | IV | 102 (84.3) | 4 (100.0) | 98 (83.8) |  | 11 (100.0) | 91 (82.7) |  |
| Histology (n, %) | Squamous cell carcinoma | 17 (14.0) | 1 (25.0) | 16 (13.7) | 1.000 | 2 (18.2) | 15 (13.6) | 1.000 |
|  | Non-squamous cell carcinoma | 104 (86.0) | 3 (75.0) | 101 (86.3) |  | 9 (81.8) | 96 (86.4) |  |
|  | Adenocarcinoma | 90 (74.4) | 2 (50.0) | 88 (75.2) |  | 7 (63.6) | 84 (75.5) |  |
|  | Large cell carcinoma | 4 (3.3) | 0 (0.0) | 4 (3.4) |  | 1 (9.1) | 3 (2.7) |  |
|  | Poorly differentiated carcinoma | 7 (5.8) | 0 (0.0) | 7 (6.0) |  | 0 (0.0) | 7 (6.4) |  |
|  | NSCLC, NOS | 3 (2.5) | 1 (25.0) | 2 (1.7) |  | 1 (9.1) | 2 (1.8) |  |
| PD-L1 IHC on TC<sup>a</sup> (n, %) | ≥50% | 18 (26.1) | 0 (0.0) | 18 (27.3) | 0.545 | 1 (14.3) | 17 (27.4) | 0.684 |
|  | 1-49% | 30 (43.5) | 1 (33.3) | 20 (30.3) |  | 2 (28.6) | 19 (30.6) |  |
|  | <1% | 21 (30.4) | 2 (66.7) | 28 (42.4) |  | 4 (57.1) | 26 (41.9) |  |
| Tumor mutational burden (n, %) | ≤10 mut/Mb | 13 (68.4) | 1 (100.0) | 12 (66.7) | 1.000 | 2 (100.0) | 11 (64.7) | 0.832 |
|  | >10 mut/Mb | 6 (31.6) | 0 (0.0) | 6 (33.3) |  | 0 (0.0) | 6 (35.3) |  |
| SLD of target lesions<sup>b</sup> (mm, mean±SD) | Pre-baseline | 61.2±37.3 | 110.8±57.4 | 59.5±35.6 | 0.006 | 75.5±52.3 | 59.7±35.4 | 0.351 |
|  | Baseline | 68.5±39.9 | 126.8±54.6 | 66.5±38.1 | 0.003 | 88.7±52.8 | 66.5±38.2 | 0.079 |
| NLR (n, %) | <5 | 64 (52.9) | 3 (75.0) | 61 (52.1) | 0.695 | 7 (63.6) | 57 (51.8) | 0.666 |
|  | ≥5 | 57 (47.1) | 1 (25.0) | 56 (47.9) |  | 4 (36.4) | 53 (48.2) |  |
| Platelets (n, %) | ≤450 (x10<sup>9</sup>/L) | 111 (91.7) | 3 (75.0) | 108 (92.3) | 0.754 | 10 (90.9) | 101 (91.8) | 1.000 |
|  | >450 (x10<sup>9</sup>/L) | 10 (8.3) | 1 (25.0) | 9 (7.7) |  | 1 (9.1) | 9 (8.2) |  |
| Treatment lines (n, %) | 1<sup>st</sup> line | 31 (25.6) | 0 (0.0) | 31 (26.5) | 0.467 | 3 (27.3) | 28 (25.5) | 0.484 |
|  | 2<sup>nd</sup> line | 63 (52.1) | 3 (75.0) | 59 (50.4) |  | 4 (36.4) | 58 (52.7) |  |
|  | ≥3<sup>rd</sup> line | 27 (22.3) | 1 (25.0) | 27 (23.1) |  | 4 (36.4) | 24 (21.8) |  |
| Type of treatment (n, %) | Immunotherapy only | 95 (78.5) | 4 (100.0) | 91 (77.8) | 0.656 | 11 (100.0) | 84 (76.4) | 0.151 |
|  | Combination with other treatments<sup>c</sup> | 26 (21.5) | 0 (0.0) | 26 (22.2) |  | 0 (0.0) | 26 (23.6) |  |
| No. of metastatic lesions (n, %) | <3 | 90 (74.4) | 2 (50.0) | 88 (75.2) | 0.580 | 6 (54.5) | 84 (76.4) | 0.223 |
|  | ≥3 | 31 (25.6) | 2 (50.0) | 29 (24.8) |  | 5 (45.5) | 26 (23.6) |  |
| Brain metastasis (n, %) | No | 89 (73.6) | 3 (75.0) | 86 (73.5) | 1.000 | 8 (72.7) | 81 (73.6) | 1.000 |
|  | Yes | 32 (26.4) | 1 (25.0) | 31 (26.5) |  | 3 (27.3) | 29 (26.4) |  |
| Bone metastasis (n, %) | No | 88 (72.7) | 1 (25.0) | 87 (74.4) | 0.108 | 6 (54.5) | 82 (74.5) | 0.287 |
|  | Yes | 33 (27.3) | 3 (75.0) | 30 (25.6) |  | 5 (45.5) | 28 (25.5) |  |
| Liver metastasis (n, %) | No | 94 (77.7) | 2 (50.0) | 92 (78.6) | 0.458 | 5 (45.5) | 89 (80.9) | 0.021 |
|  | Yes | 27 (22.3) | 2 (50.0) | 25 (21.4) |  | 6 (54.5) | 21 (19.1) |  |

<sup>a</sup>PD-L1 results were available only in 69 (57.0%) patients.; <sup>b</sup>SLD of target lesions was measured based on RECIST 1.1.; <sup>c</sup>Other treatments were chemotherapy (n=22), targeted therapy (n=2), and clinical trial drugs (n=2).
NOS, not other specified; IHC, immunohistochemistry; TC, tumor cells; PS, performance status; Mut/Mb, mutation per megabase; NLR, neutrophil-to-lymphocyte ratio; HPDc, hyperprogression according to the definition proposed by *Champrat* et al [8]; HPDs, hyperprogression according to the definition proposed by *Saâda-Bouzid* et al [10].

**Supplementary Table 3.** Target lesions and changes of tumor size at each time point based on RECIST 1.1 among patients with HPD or mHPD

| No. of case | Target lesions | Pre-baseline (mm) | Baseline (mm) | Post-treatment (mm) | Follow-up CT after 3 months (mm) | HPD c | mHPDc | HPD s | mHPD s | HPDf | mHPDf |
| --- | --- | --- | --- | --- | --- | --- | --- | --- | --- | --- | --- |
| 1 | Para-aortic lymph node | 11 | 22 | 26 | N/A | Y | Y | Y | Y | N | Y |
|  | Right chest soft tissue | 42 | 52 | 83 |  |  |  |  |  |  |  |
|  | Left lower lobe mass | 35 | 49 | 70 |  |  |  |  |  |  |  |
|  | Right axillary lymph node |  |  | 18 |  |  |  |  |  |  |  |
|  | Inferior right hilar lymph node |  |  | 20 |  |  |  |  |  |  |  |
|  | Left supraclavicular lymph node |  |  | 17 |  |  |  |  |  |  |  |
|  | New mass on a major fissure |  |  | 40 |  |  |  |  |  |  |  |
| 2 | Hepatic dome mass | 12 | 17 | 23 | N/A | Y | Y | Y | Y | N | Y |
|  | Left hepatic lobe mass | 17 | 19 | 22 |  |  |  |  |  |  |  |
|  | Left upper lobe mass | 37 | 39 | 43 |  |  |  |  |  |  |  |
|  | Left pedicle lytic lesion |  |  | 26 |  |  |  |  |  |  |  |
|  | Lytic lesion right sternum |  |  | 15 |  |  |  |  |  |  |  |
|  | Upper pole right kidney |  |  | 25 |  |  |  |  |  |  |  |
|  | Upper pole left kidney |  |  | 21 |  |  |  |  |  |  |  |
| 3 | Right paratracheal nodule | 20 | 23 | 20 | 21 | Y | Y | Y | Y | N | N |
|  | Retrocrural mass | 50 | 57 | 89 | 103 |  |  |  |  |  |  |
|  | Liver lesion | 62 | 63 | 63 | 56 |  |  |  |  |  |  |
|  | Left adrenal soft tissue | 63 | 60 | 79 | 86 |  |  |  |  |  |  |
| 4 | Left upper lobe mass | 73 | 77 | 122 | N/A | Y | Y | Y | Y | N | Y |
|  | Left lower lobe nodule | 10 | 16 | 5 |  |  |  |  |  |  |  |
|  | Right lobe liver mass | 11 | 13 | 13 |  |  |  |  |  |  |  |
|  | Left hilar lymph node |  |  | 15 |  |  |  |  |  |  |  |
|  | Destructive osseous metastases |  |  | 16 |  |  |  |  |  |  |  |
| 5 | Posterior right paratracheal mass | 5 | 30 | 93 | N/A | N | Y | Y | Y | N | Y |
| 6 | Right lower lobe | 14 | 18 | 21 | N/A | N | N | Y | Y | N | N |
| 7 | Left upper lobe mass | 45 | 45 | 70 | N/A | N | Y | Y | Y | N | Y |
|  | Left apex mass | 28 | 28 | 40 |  |  |  |  |  |  |  |
|  | Left subcarinal lymph node | 10 | 18 | 14 |  |  |  |  |  |  |  |
|  | Destructive metastatic lesion in left scapula |  |  | 64 |  |  |  |  |  |  |  |
| 8 | Right lower paratracheal node | 20 | 28 | 88 | 51 | N | Y | Y | Y | N | N |
|  | Subcarinal lymph node | 15 | 30 | 46 | 69 |  |  |  |  |  |  |
|  | Right upper lobe mass | 33 | 40 | 52 | 44 |  |  |  |  |  |  |
|  | Right pleural nodule 1 |  |  | 27 | 25 |  |  |  |  |  |  |
|  | Right pleural nodule 2 |  |  | 18 | 14 |  |  |  |  |  |  |
|  | Right upper lobe node |  |  | 19 | 32 |  |  |  |  |  |  |
|  | Right hepatic lobe mass |  |  | 18 | 21 |  |  |  |  |  |  |
|  | Hepatic dome mass |  |  | 15 | 15 |  |  |  |  |  |  |
|  | Caudal liver mass |  |  | 26 | 26 |  |  |  |  |  |  |
| 9 | Posterior mediastinal mass | 87 | 88 | 92 | N/A | N | N | Y | Y | N | N |
| 10 | Right lung nodule | 26 | 27 | 30 | 45 | N | N | Y | Y | N | N |
| 11 | Medial right lung | 20 | 27 | 27 | 31 | N | N | Y | Y | N | N |
|  | Right pleural nodule | 10 | 17 | 23 | 39 |  |  |  |  |  |  |
|  | Right supraclavicular soft tissue | 55 | 45 | 58 | 60 |  |  |  |  |  |  |
|  | Basilar nodule | 17 | 18 | 18 | 20 |  |  |  |  |  |  |
|  | Left lower lobe nodule | 7 | 10 | 11 | 15 |  |  |  |  |  |  |
| 12 | Lung nodule in right upper lobe | 8 | 10 | 8 | 7 | N | Y | N | Y | N | Y |
|  | Thyroid mass in left lobe | 23 | 29 | 29 | 28 |  |  |  |  |  |  |
|  | Mass in right costo-vertebral junction | 29 | 34 | 35 | 35 |  |  |  |  |  |  |
|  | Vertebral body metastatic |  |  | 38 | 52 |  |  |  |  |  |  |
|  | Hepatic mass in left lobe |  |  | 15 | 16 |  |  |  |  |  |  |
| 13 | Right paratracheal lymph node | 22 | 38 | 38 | N/A<sup>a</sup> | N | Y | N | Y | N | Y |
|  | Right upper lobe nodule | 13 | 15 | 16 |  |  |  |  |  |  |  |
|  | Right middle lobe nodule | 33 | 35 | 35 |  |  |  |  |  |  |  |
|  | Fibrosis mass in the anterior right upper lobe |  |  | 39 |  |  |  |  |  |  |  |
|  | Subcarinal lymph node |  |  | 35 |  |  |  |  |  |  |  |
|  | Right supraclavicular lymph node |  |  | 18 |  |  |  |  |  |  |  |
|  | Right paraspinal muscle nodule |  |  | 19 |  |  |  |  |  |  |  |
|  | Right adrenal nodule |  |  | 17 |  |  |  |  |  |  |  |
|  | Left adrenal nodule |  |  | 30 |  |  |  |  |  |  |  |
|  | Gastrohepatic lymph node |  |  | 39 |  |  |  |  |  |  |  |
|  | Soft tissue lesion on right kidney |  |  | 40 |  |  |  |  |  |  |  |
|  | Soft tissue left kidney |  |  | 26 |  |  |  |  |  |  |  |
| 14 | Hilar soft tissue | 34 | 36 | 36 | 11 | N | Y | N | Y | N | Y |
|  | Left adrenal nodule 1 | 18 | 18 | 19 | 19 |  |  |  |  |  |  |
|  | Left adrenal nodule 2 | 8 | 12 | 12 | 19 |  |  |  |  |  |  |
|  | Posterior omental node |  |  | 17 | 35 |  |  |  |  |  |  |
| 15 | Lung mass | 89 | 100 | 125 | N/A<sup>a</sup> | N | Y | N | Y | N | N |
|  | Posterior right hepatic lobe |  |  | 28 |  |  |  |  |  |  |  |
|  | Right middle lobe node |  |  | 19 |  |  |  |  |  |  |  |
|  | Left lower lobe node |  |  | 19 |  |  |  |  |  |  |  |
| 16 | Right upper lobe mass | 21 | 28 | 38 | 49 | N | Y | N | Y | N | Y |
|  | Right lower lobe mass | 17 | 23 | 32 | 43 |  |  |  |  |  |  |
|  | Subcarinal lymph node |  |  | 14 | 14 |  |  |  |  |  |  |
|  | Right axis hilar lymph node |  |  | 14 | 26 |  |  |  |  |  |  |
|  | Pancreatic hepatic lymph node |  |  | 16 | 16 |  |  |  |  |  |  |
| 17 | Right upper lobe nodule | 4 | 11 | 16 | N/A<sup>a</sup> | N | Y | N | Y | N | Y |
|  | Left upper lobe nodule |  |  | 11 |  |  |  |  |  |  |  |
|  | Right paratracheal lymph node |  |  | 22 |  |  |  |  |  |  |  |
| 18 | Right hilar lymph node | 22 | 22 | 24 | N/A<sup>a</sup> | N | Y | N | N | N | Y |
|  | Pleural nodule in left apex |  | 23 | 28 |  |  |  |  |  |  |  |
|  | Lung mass at right upper lobe |  | 19 | 30 |  |  |  |  |  |  |  |
|  | Left hilar lymph node |  |  | 16 |  |  |  |  |  |  |  |
| 19 | Right hilar lymph node | 22 | 22 | 24 | N/A<sup>a</sup> | N | N | N | N | N | Y |
|  | Left apical pleural nodule |  | 23 | 28 |  |  |  |  |  |  |  |
|  | Right upper lobe mass |  | 19 | 30 |  |  |  |  |  |  |  |
|  | Left hilar lymph node |  |  | 16 |  |  |  |  |  |  |  |

HPDc, HPDs, and HPDf, HPD according to the definition proposed by *Champiat et al* [8], *Saâda-Bouزيد et al* [10] and *Ferrara et al* [11].; mHPDc and mHPDs, HPD according to *Champiat et al* and *Saâda-Bouزيد et al.* along with the inclusion of new measurable lesions seen at the post-treatment CT scan.

**Supplementary Table 4.**
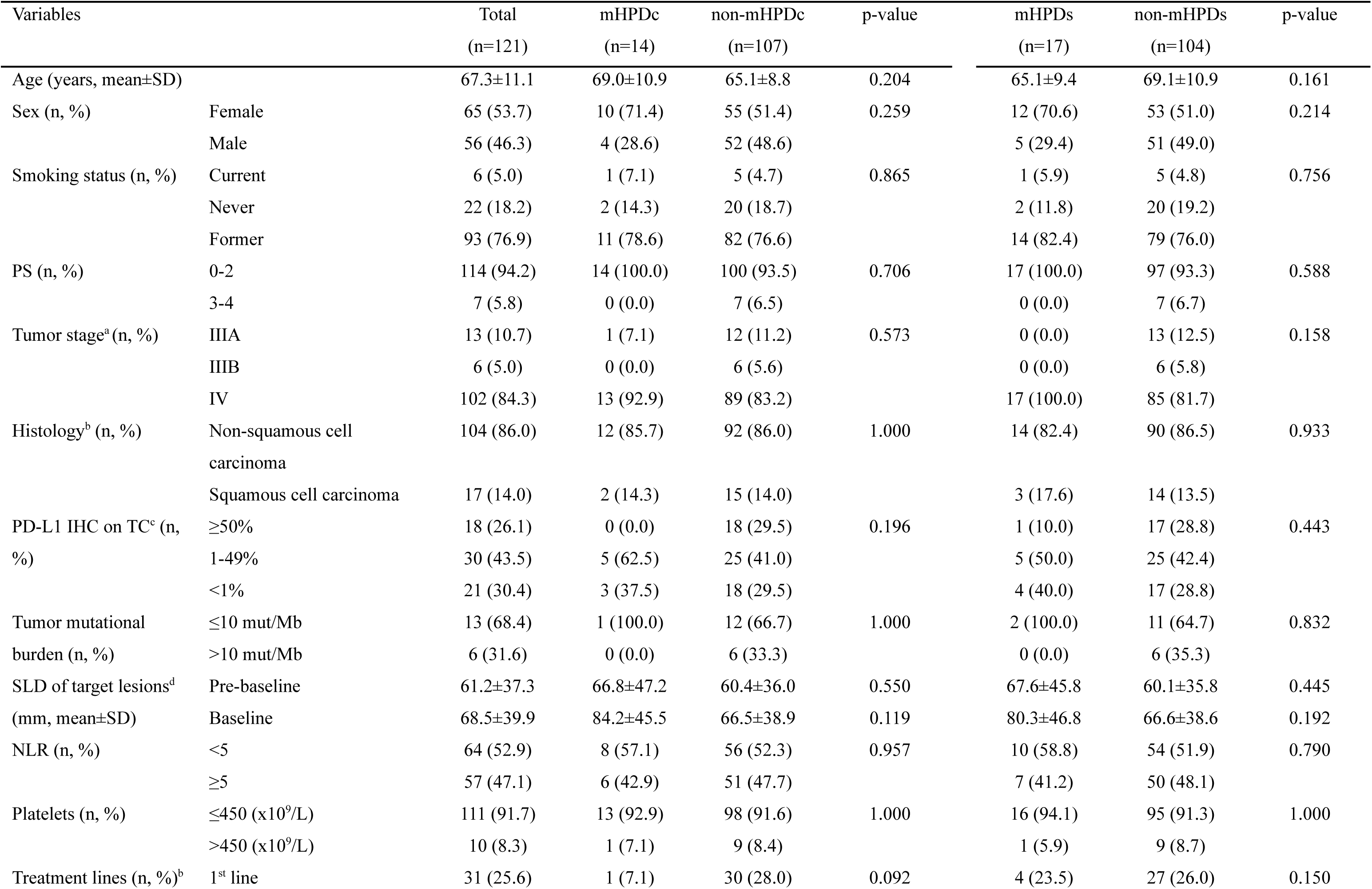
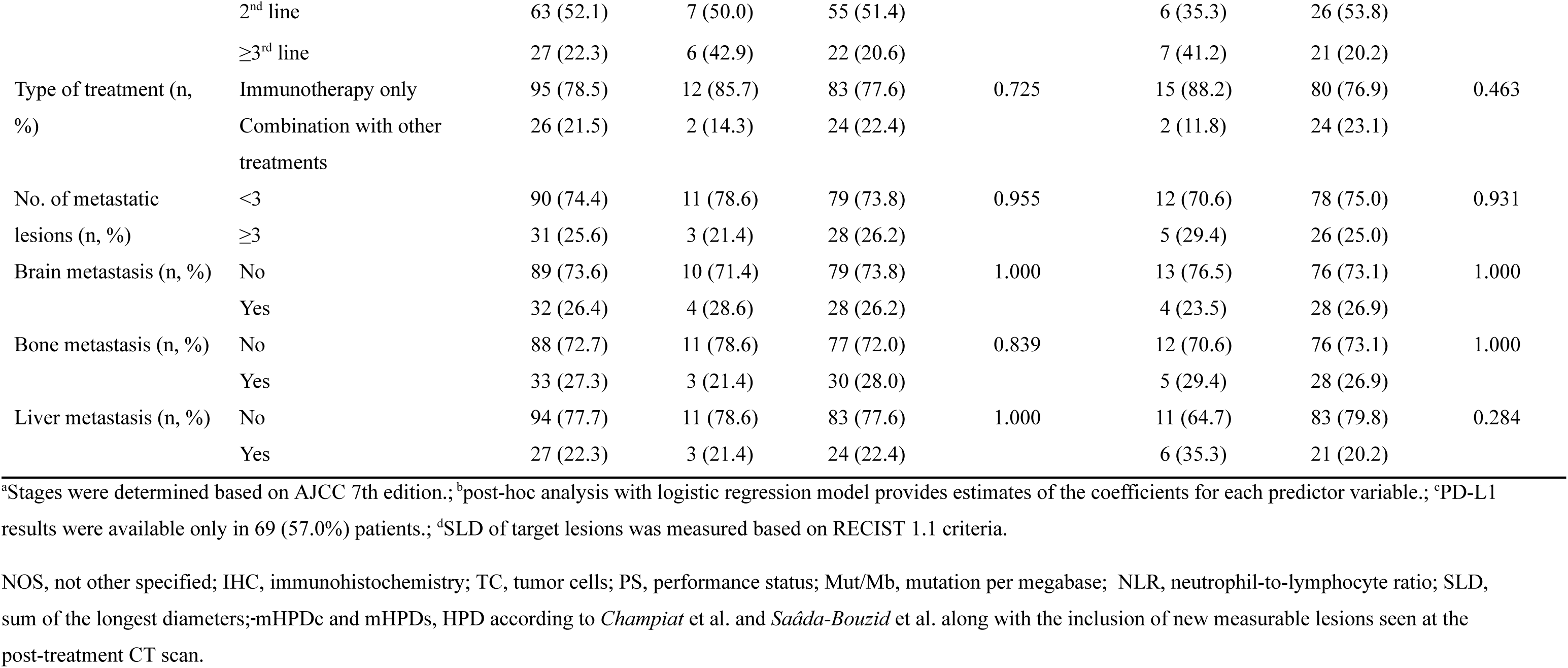
Clinical and pathological characteristics between patients with and without hyperprogressive disease (HPD) based on each modified HPD

| Variables |  | Total<br>(n=121) | mHPDc<br>(n=14) | non-mHPDc<br>(n=107) | p-value | mHPDs<br>(n=17) | non-mHPDs<br>(n=104) | p-value |
| --- | --- | --- | --- | --- | --- | --- | --- | --- |
| Age (years, mean±SD) |  | 67.3±11.1 | 69.0±10.9 | 65.1±8.8 | 0.204 | 65.1±9.4 | 69.1±10.9 | 0.161 |
| Sex (n, %) | Female | 65 (53.7) | 10 (71.4) | 55 (51.4) | 0.259 | 12 (70.6) | 53 (51.0) | 0.214 |
|  | Male | 56 (46.3) | 4 (28.6) | 52 (48.6) |  | 5 (29.4) | 51 (49.0) |  |
| Smoking status (n, %) | Current | 6 (5.0) | 1 (7.1) | 5 (4.7) | 0.865 | 1 (5.9) | 5 (4.8) | 0.756 |
|  | Never | 22 (18.2) | 2 (14.3) | 20 (18.7) |  | 2 (11.8) | 20 (19.2) |  |
|  | Former | 93 (76.9) | 11 (78.6) | 82 (76.6) |  | 14 (82.4) | 79 (76.0) |  |
| PS (n, %) | 0-2 | 114 (94.2) | 14 (100.0) | 100 (93.5) | 0.706 | 17 (100.0) | 97 (93.3) | 0.588 |
|  | 3-4 | 7 (5.8) | 0 (0.0) | 7 (6.5) |  | 0 (0.0) | 7 (6.7) |  |
| Tumor stage<sup>a</sup> (n, %) | IIIA | 13 (10.7) | 1 (7.1) | 12 (11.2) | 0.573 | 0 (0.0) | 13 (12.5) | 0.158 |
|  | IIIB | 6 (5.0) | 0 (0.0) | 6 (5.6) |  | 0 (0.0) | 6 (5.8) |  |
|  | IV | 102 (84.3) | 13 (92.9) | 89 (83.2) |  | 17 (100.0) | 85 (81.7) |  |
| Histology<sup>b</sup> (n, %) | Non-squamous cell carcinoma | 104 (86.0) | 12 (85.7) | 92 (86.0) | 1.000 | 14 (82.4) | 90 (86.5) | 0.933 |
|  | Squamous cell carcinoma | 17 (14.0) | 2 (14.3) | 15 (14.0) |  | 3 (17.6) | 14 (13.5) |  |
| PD-L1 IHC on TC<sup>c</sup> (n, %) | ≥50% | 18 (26.1) | 0 (0.0) | 18 (29.5) | 0.196 | 1 (10.0) | 17 (28.8) | 0.443 |
|  | 1-49% | 30 (43.5) | 5 (62.5) | 25 (41.0) |  | 5 (50.0) | 25 (42.4) |  |
|  | <1% | 21 (30.4) | 3 (37.5) | 18 (29.5) |  | 4 (40.0) | 17 (28.8) |  |
| Tumor mutational burden (n, %) | ≤10 mut/Mb | 13 (68.4) | 1 (100.0) | 12 (66.7) | 1.000 | 2 (100.0) | 11 (64.7) | 0.832 |
|  | >10 mut/Mb | 6 (31.6) | 0 (0.0) | 6 (33.3) |  | 0 (0.0) | 6 (35.3) |  |
| SLD of target lesions<sup>d</sup> (mm, mean±SD) | Pre-baseline | 61.2±37.3 | 66.8±47.2 | 60.4±36.0 | 0.550 | 67.6±45.8 | 60.1±35.8 | 0.445 |
|  | Baseline | 68.5±39.9 | 84.2±45.5 | 66.5±38.9 | 0.119 | 80.3±46.8 | 66.6±38.6 | 0.192 |
| NLR (n, %) | <5 | 64 (52.9) | 8 (57.1) | 56 (52.3) | 0.957 | 10 (58.8) | 54 (51.9) | 0.790 |
|  | ≥5 | 57 (47.1) | 6 (42.9) | 51 (47.7) |  | 7 (41.2) | 50 (48.1) |  |
| Platelets (n, %) | ≤450 (x10<sup>9</sup>/L) | 111 (91.7) | 13 (92.9) | 98 (91.6) | 1.000 | 16 (94.1) | 95 (91.3) | 1.000 |
|  | >450 (x10<sup>9</sup>/L) | 10 (8.3) | 1 (7.1) | 9 (8.4) |  | 1 (5.9) | 9 (8.7) |  |
| Treatment lines (n, %)<sup>b</sup> | 1<sup>st</sup> line | 31 (25.6) | 1 (7.1) | 30 (28.0) | 0.092 | 4 (23.5) | 27 (26.0) | 0.150 |
|  | 2<sup>nd</sup> line | 63 (52.1) | 7 (50.0) | 55 (51.4) |  | 6 (35.3) | 26 (53.8) |  |
|  | ≥3<sup>rd</sup> line | 27 (22.3) | 6 (42.9) | 22 (20.6) |  | 7 (41.2) | 21 (20.2) |  |
| Type of treatment (n, %) | Immunotherapy only | 95 (78.5) | 12 (85.7) | 83 (77.6) | 0.725 | 15 (88.2) | 80 (76.9) | 0.463 |
|  | Combination with other treatments | 26 (21.5) | 2 (14.3) | 24 (22.4) |  | 2 (11.8) | 24 (23.1) |  |
| No. of metastatic lesions (n, %) | <3 | 90 (74.4) | 11 (78.6) | 79 (73.8) | 0.955 | 12 (70.6) | 78 (75.0) | 0.931 |
|  | ≥3 | 31 (25.6) | 3 (21.4) | 28 (26.2) |  | 5 (29.4) | 26 (25.0) |  |
| Brain metastasis (n, %) | No | 89 (73.6) | 10 (71.4) | 79 (73.8) | 1.000 | 13 (76.5) | 76 (73.1) | 1.000 |
|  | Yes | 32 (26.4) | 4 (28.6) | 28 (26.2) |  | 4 (23.5) | 28 (26.9) |  |
| Bone metastasis (n, %) | No | 88 (72.7) | 11 (78.6) | 77 (72.0) | 0.839 | 12 (70.6) | 76 (73.1) | 1.000 |
|  | Yes | 33 (27.3) | 3 (21.4) | 30 (28.0) |  | 5 (29.4) | 28 (26.9) |  |
| Liver metastasis (n, %) | No | 94 (77.7) | 11 (78.6) | 83 (77.6) | 1.000 | 11 (64.7) | 83 (79.8) | 0.284 |
|  | Yes | 27 (22.3) | 3 (21.4) | 24 (22.4) |  | 6 (35.3) | 21 (20.2) |  |

<sup>a</sup>Stages were determined based on AJCC 7th edition.; <sup>b</sup>post-hoc analysis with logistic regression model provides estimates of the coefficients for each predictor variable.; <sup>c</sup>PD-L1 results were available only in 69 (57.0%) patients.; <sup>d</sup>SLD of target lesions was measured based on RECIST 1.1 criteria.
NOS, not other specified; IHC, immunohistochemistry; TC, tumor cells; PS, performance status; Mut/Mb, mutation per megabase; NLR, neutrophil-to-lymphocyte ratio; SLD, sum of the longest diameters; mHPDc and mHPDs, HPD according to *Champiat* et al. and *Saâda-Bouزيد* et al. along with the inclusion of new measurable lesions seen at the post-treatment CT scan.

**Supplementary Table 5.**
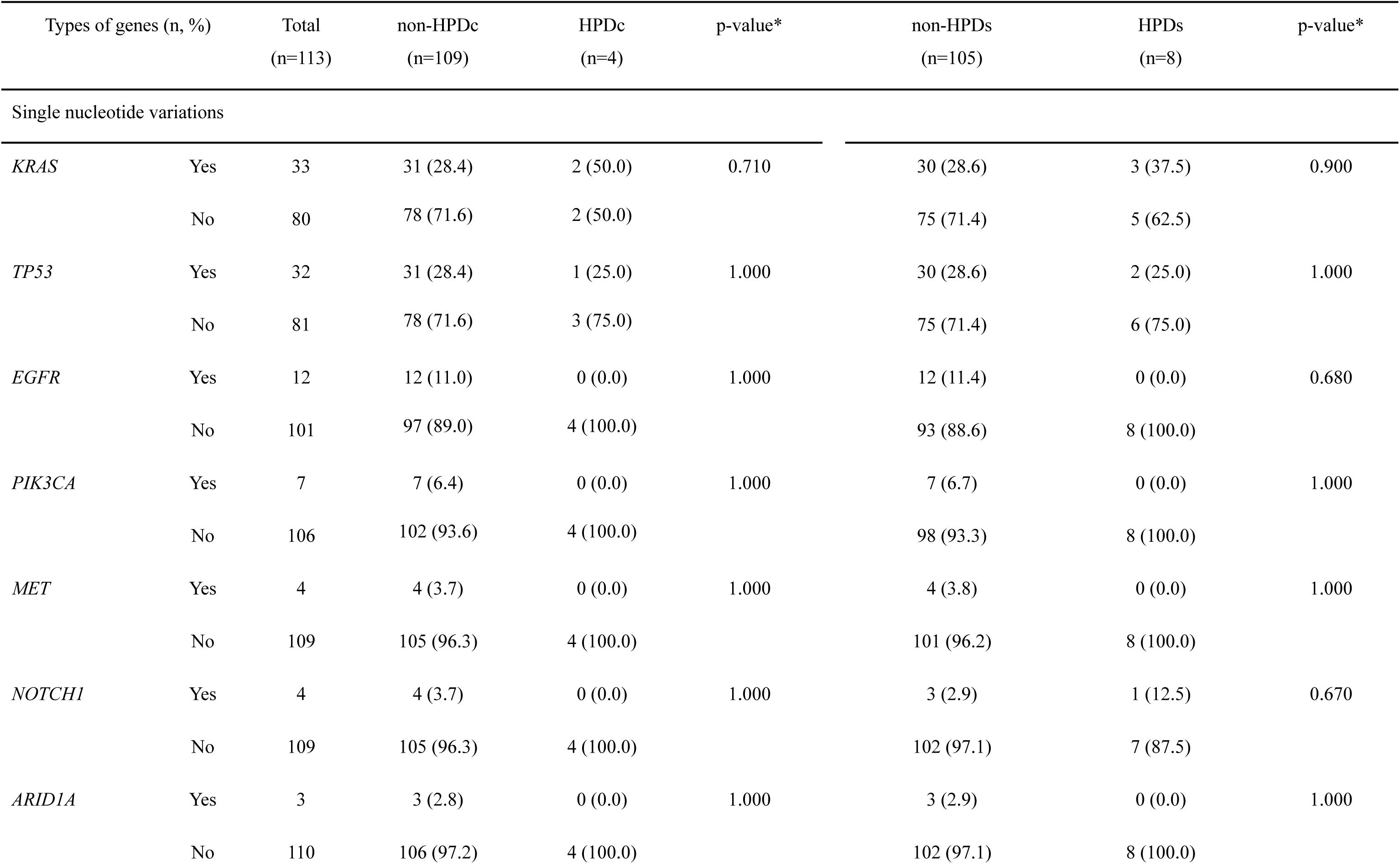
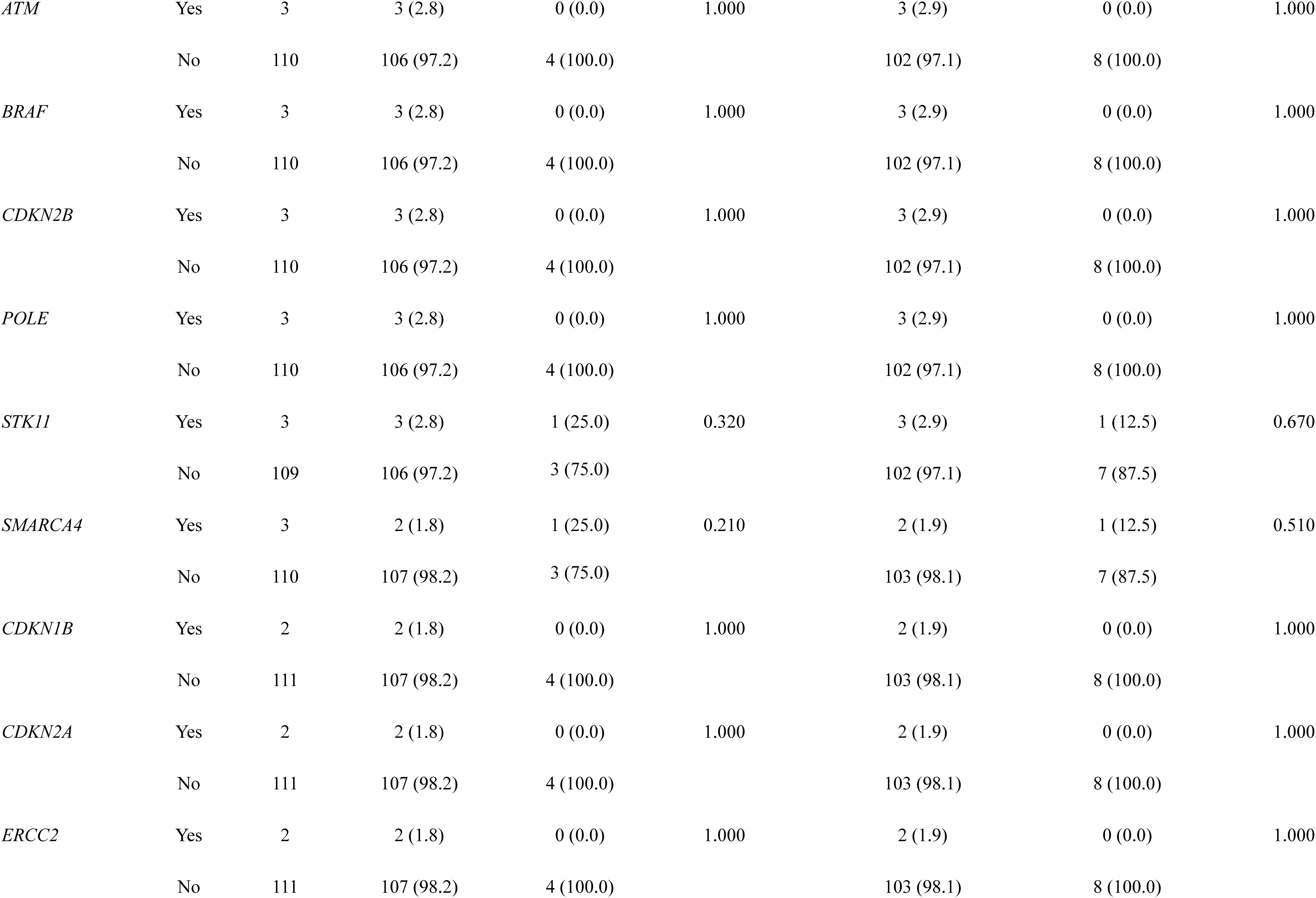
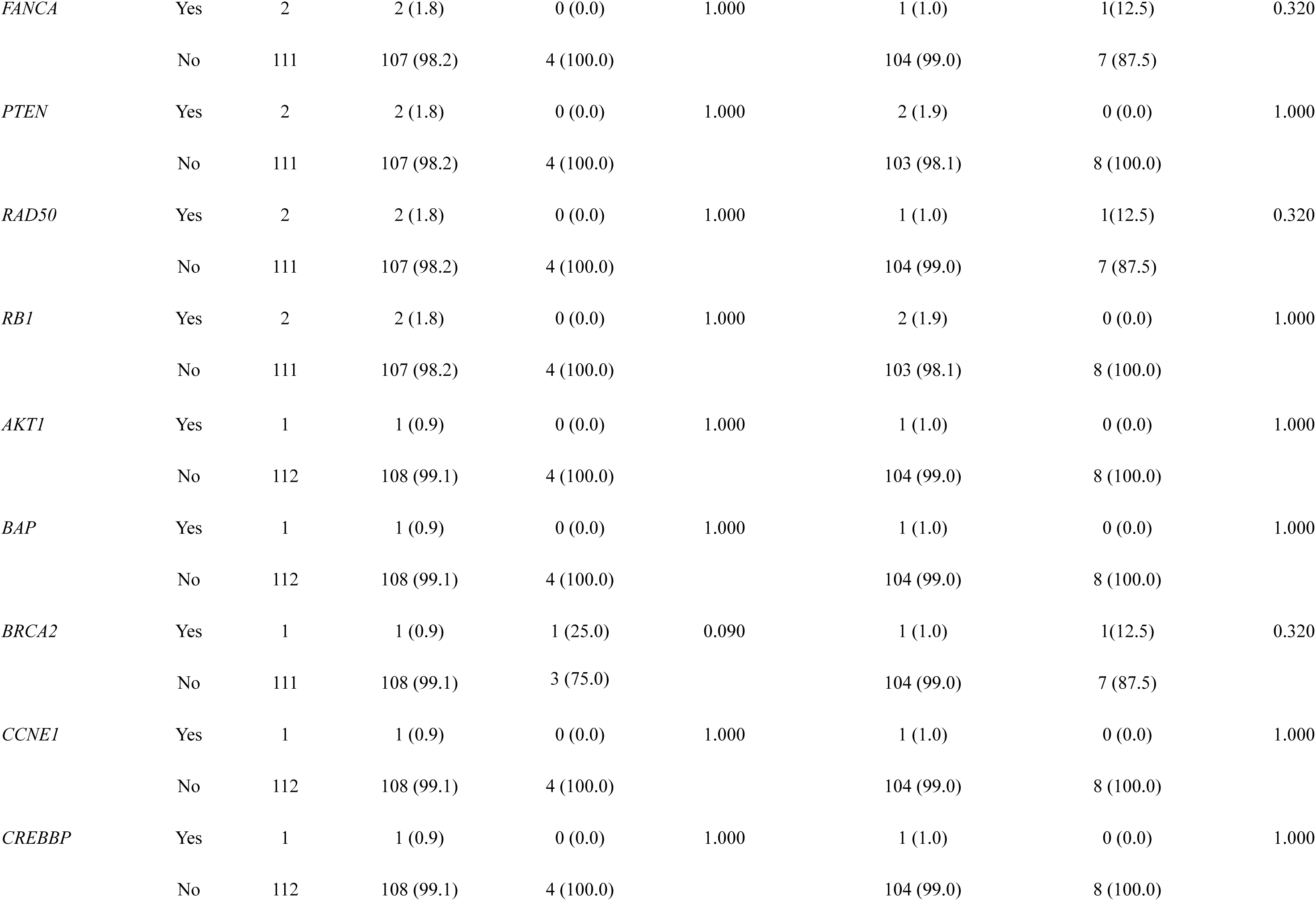
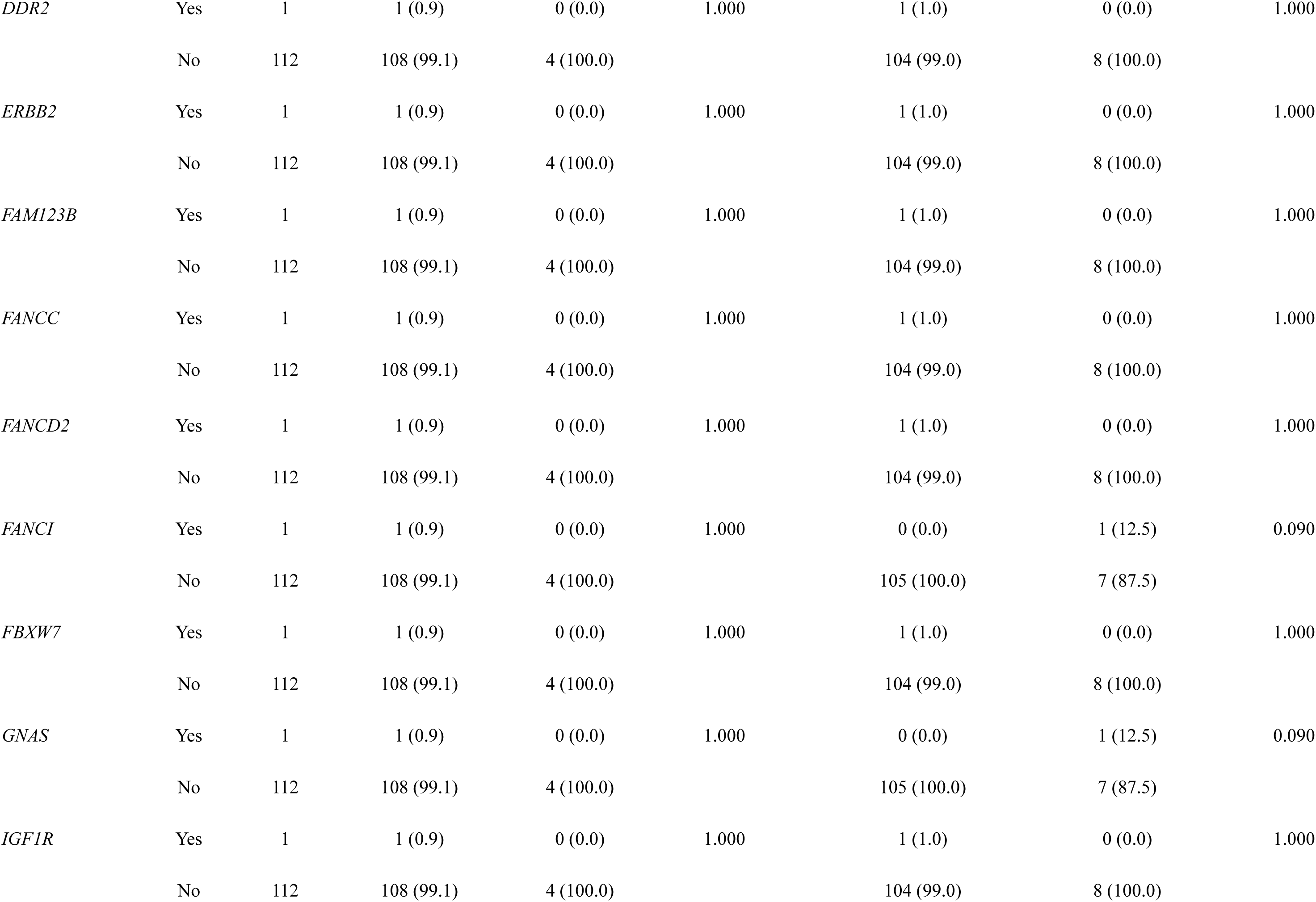
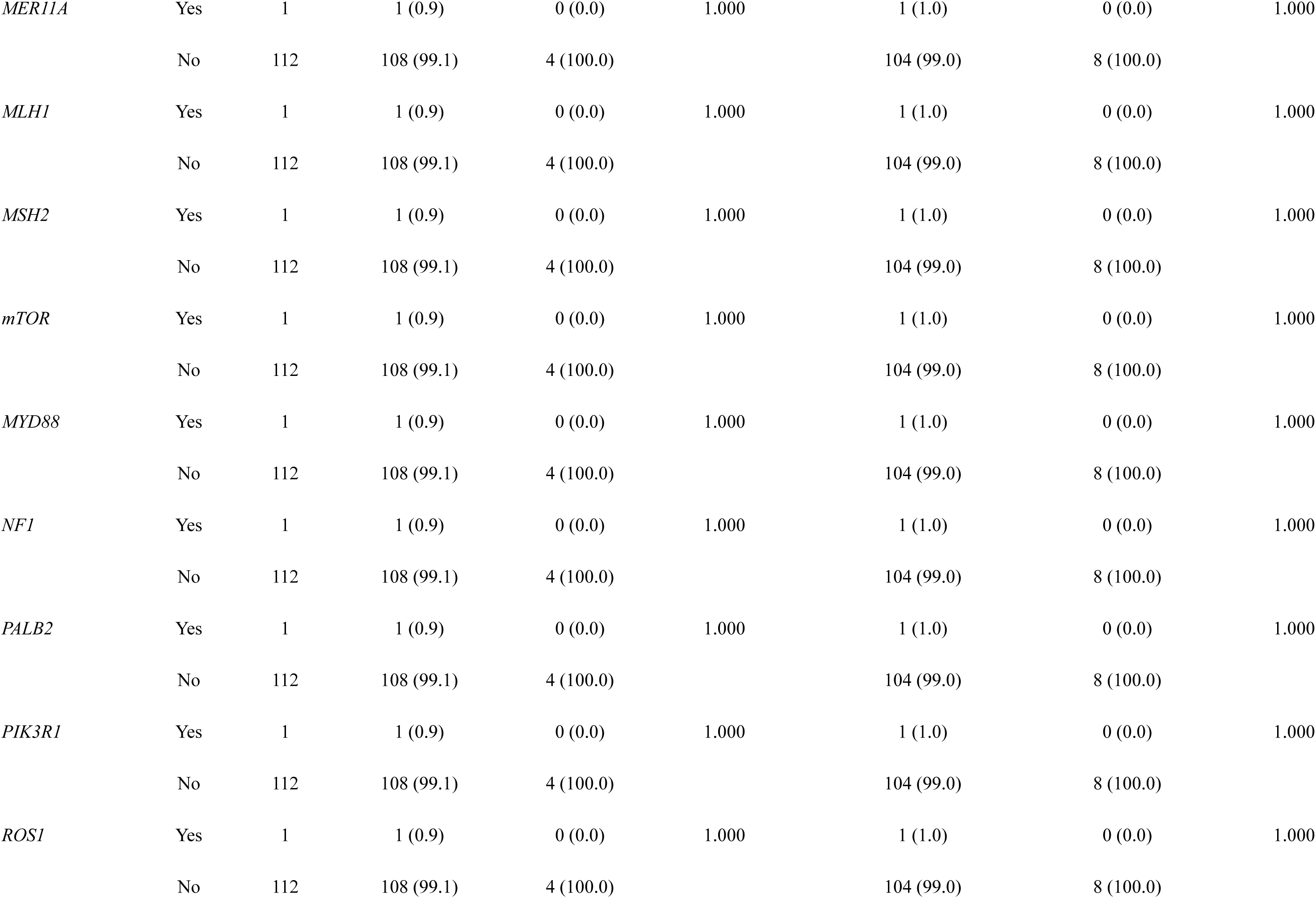
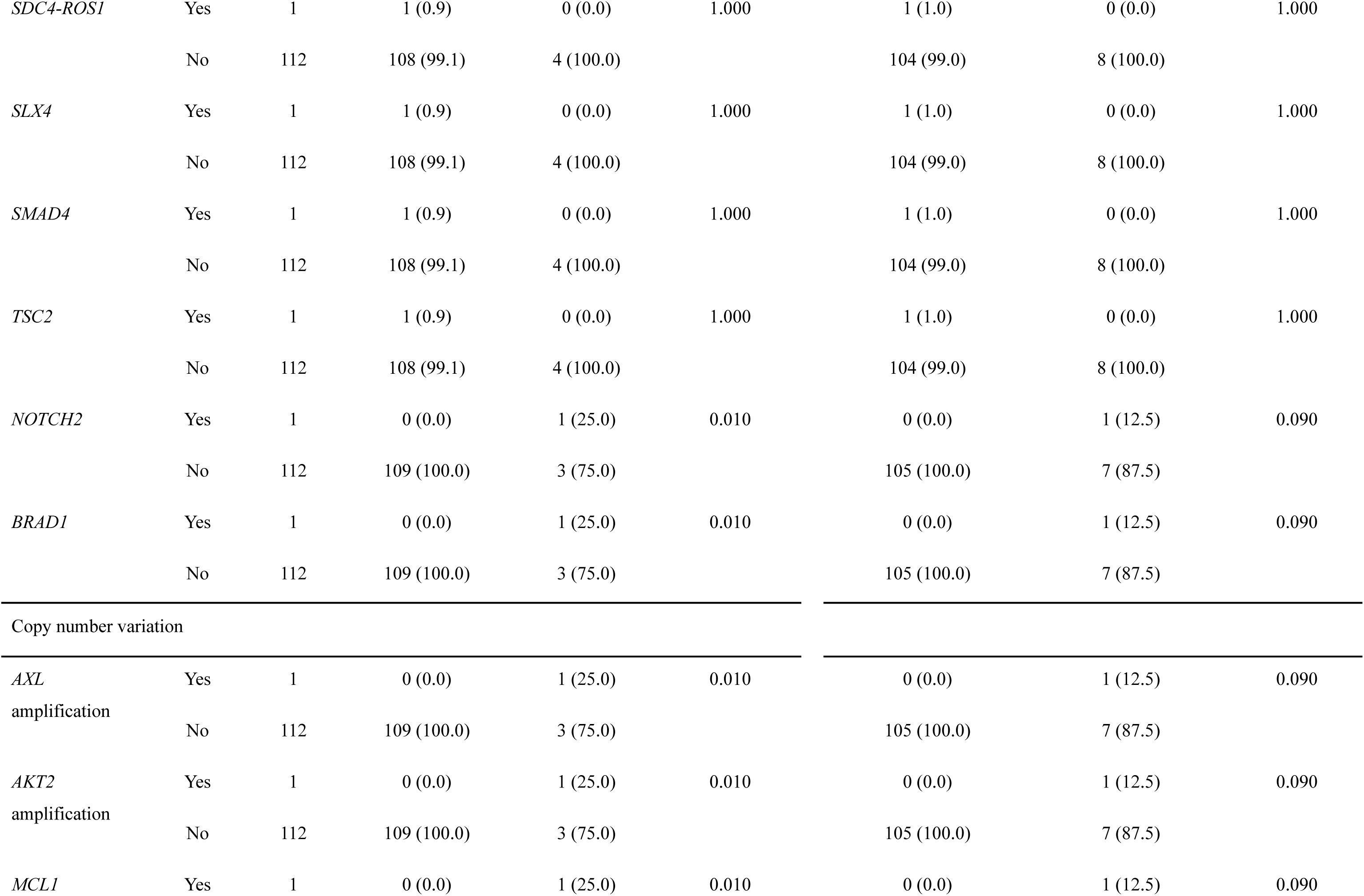
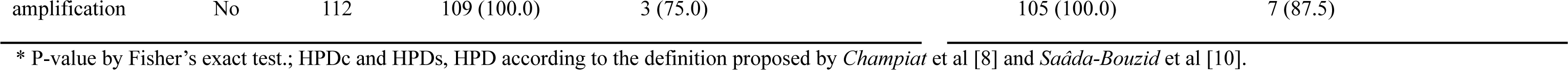
Genomic alterations between patients with and without hyperprogressive disease (HPD) based on each definition of HPD

## Reference

1. Felip E, Altorki N, Zhou C, Csőszi T, Vynnychenko I, Goloborodko O, et al. Adjuvant atezolizumab after adjuvant chemotherapy in resected stage IB-IIIA non-small-cell lung cancer (IMpower010): a randomised, multicentre, open-label, phase 3 trial. Lancet. 2021;398(10308):1344–57. Epub 2021/09/24. doi: 10.1016/s0140-6736(21)02098-5. PubMed PMID: 34555333.

2. Mok TSK, Wu YL, Kudaba I, Kowalski DM, Cho BC, Turna HZ, et al. Pembrolizumab versus chemotherapy for previously untreated, PD-L1-expressing, locally advanced or metastatic non-small-cell lung cancer (KEYNOTE-042): a randomised, open-label, controlled, phase 3 trial. Lancet. 2019;393(10183):1819–30. Epub 2019/04/09. doi: 10.1016/s0140-6736(18)32409-7. PubMed PMID: 30955977.

3. Paz-Ares L, Ciuleanu TE, Cobo M, Schenker M, Zurawski B, Menezes J, et al. First-line nivolumab plus ipilimumab combined with two cycles of chemotherapy in patients with non-small-cell lung cancer (CheckMate 9LA): an international, randomised, open-label, phase 3 trial. Lancet Oncol. 2021;22(2):198–211. Epub 2021/01/22. doi: 10.1016/s1470-2045(20)30641-0. PubMed PMID: 33476593.

4. Borcoman E, Kanjanapan Y, Champiat S, Kato S, Servois V, Kurzrock R, et al. Novel patterns of response under immunotherapy. Ann Oncol. 2019;30(3):385–96. Epub 2019/01/19. doi: 10.1093/annonc/mdz003. PubMed PMID: 30657859.

5. Zhou L, Zhang M, Li R, Xue J, Lu Y. Pseudoprogression and hyperprogression in lung cancer: a comprehensive review of literature. J Cancer Res Clin Oncol. 2020;146(12):3269–79. Epub 2020/08/29. doi: 10.1007/s00432-020-03360-1. PubMed PMID: 32857178.

6. Kurman JS, Murgu SD. Hyperprogressive disease in patients with non-small cell lung cancer on immunotherapy. J Thorac Dis. 2018;10(2):1124–8. Epub 2018/04/03. doi: 10.21037/jtd.2018.01.79. PubMed PMID: 29607190; PubMed Central PMCID: PMCPMC5864602.

7. Abbar B, De Castelbajac V, Gougis P, Assoun S, Pluvy J, Tesmoingt C, et al. Definitions, outcomes, and management of hyperprogression in patients with non-small-cell lung cancer treated with immune checkpoint inhibitors. Lung Cancer. 2021;152:109–18. Epub 2021/01/02. doi: 10.1016/j.lungcan.2020.12.026. PubMed PMID: 33385736.

8. Champiat S, Dercle L, Ammari S, Massard C, Hollebecque A, Postel-Vinay S, et al. Hyperprogressive Disease Is a New Pattern of Progression in Cancer Patients Treated by Anti-PD-1/PD-L1. Clin Cancer Res. 2017;23(8):1920–8. Epub 2016/11/09. doi: 10.1158/1078-0432.Ccr-16-1741. PubMed PMID: 27827313.

9. Kim SH, Choi CM, Lee DH, Kim SW, Yoon S, Kim WS, et al. Clinical outcomes of nivolumab in patients with advanced non-small cell lung cancer in real-world practice, with an emphasis on hyper-progressive disease. J Cancer Res Clin Oncol. 2020;146(11):3025–36. Epub 2020/06/26. doi: 10.1007/s00432-020-03293-9. PubMed PMID: 32583235.

10. Saâda-Bouzid E, Defaucheux C, Karabajakian A, Coloma VP, Servois V, Paoletti X, et al. Hyperprogression during anti-PD-1/PD-L1 therapy in patients with recurrent and/or metastatic head and neck squamous cell carcinoma. Ann Oncol. 2017;28(7):1605–11. Epub 2017/04/19. doi: 10.1093/annonc/mdx178. PubMed PMID: 28419181.

11. Ferrara R, Mezquita L, Texier M, Lahmar J, Audigier-Valette C, Tessonnier L, et al. Hyperprogressive Disease in Patients With Advanced Non-Small Cell Lung Cancer Treated With PD-1/PD-L1 Inhibitors or With Single-Agent Chemotherapy. JAMA Oncol. 2018;4(11):1543–52. Epub 2018/09/08. doi: 10.1001/jamaoncol.2018.3676. PubMed PMID: 30193240; PubMed Central PMCID: PMCPMC6248085 consulting/advisory role for Bristol-Myers Squibb and Merck Sharp & Dohme. Dr Mazieres reported serving in a consulting/advisory role for Bristol-Myers Squibb, Merck Sharp & Dohme, Roche, AstraZeneca, and Eli Lilly and reported receiving research funding from Roche, Bristol-Myers Squibb, and AstraZeneca. Dr Duchemann reported serving in a consulting/advisory role for Roche and Bristol-Myers Squibb and reported receiving travel/accommodation funding from Roche. Dr Westeel reported serving in a consulting/advisory role for Bristol-Myers Squibb, Merck Sharp & Dohme, and AstraZeneca; reported receiving honoraria from Bristol-Myers Squibb and AstraZeneca; reported serving on speakers bureaus for Bristol-Myers Squibb and Merck Sharp & Dohme; and reported receiving travel/accommodation funding from Bristol-Myers Squibb, Roche, and AstraZeneca. Dr Remon reported receiving travel/accommodation funding from Merck Sharp & Dohme and OSE Pharma. Dr Adam reported serving in a consulting/advisory role for Roche, Bristol-Myers Squibb, Merck Sharp & Dohme, and AstraZeneca. Dr Bria reported receiving speakers fees from Bristol-Myers Squibb, Novartis, AstraZeneca, Merck Sharp & Dohme, Celgene, Pfizer, Helsinn, Eli Lilly, and Roche and reported receiving research funding from AstraZeneca, Roche, Open Innovation, Italian Association for Cancer Research (AIRC), and Cariverona Foundation. Dr Soria reported serving in a consulting/advisory role for Roche, AstraZeneca, and Pfizer. Dr Besse reported receiving research funding from GlaxoSmithKline, Roche/Genentech, Clovis Oncology, Pfizer, Boehringer, Eli Lilly, Servier, Onxeo, Bristol-Myers Squibb, Merck Sharp & Dohme, OSE Pharma, Inivata, and AstraZeneca. No other disclosures were reported.

12. Kato S, Goodman A, Walavalkar V, Barkauskas DA, Sharabi A, Kurzrock R. Hyperprogressors after Immunotherapy: Analysis of Genomic Alterations Associated with Accelerated Growth Rate. Clin Cancer Res. 2017;23(15):4242–50. Epub 2017/03/30. doi: 10.1158/1078-0432.Ccr-16-3133. PubMed PMID: 28351930; PubMed Central PMCID: PMCPMC5647162.

13. Singavi AK, Menon S, Kilari D, Alqwasmi A, Ritch PS, Thomas JP, et al. Predictive biomarkers for hyper-progression (HP) in response to immune checkpoint inhibitors (ICI)-analysis of somatic alterations (SAs). Annals of Oncology. 2017;28:v405. doi: 10.1093/annonc/mdx376.006.

14. Wang M, Huang H, Xu Z, Li Z, Shen L, Yu Y, et al. Proposal for multiple new lesions as complement of hyperprogressive disease in NSCLC patients treated with PD-1/PD-L1 immunotherapy. Lung Cancer. 2022;173:28–34. Epub 2022/09/19. doi: 10.1016/j.lungcan.2022.09.001. PubMed PMID: 36116167.

15. Champiat S, Ferrara R, Massard C, Besse B, Marabelle A, Soria JC, et al. Hyperprogressive disease: recognizing a novel pattern to improve patient management. Nat Rev Clin Oncol. 2018;15(12):748–62. Epub 2018/10/27. doi: 10.1038/s41571-018-0111-2. PubMed PMID: 30361681.

16. Bagley SJ, Kothari S, Aggarwal C, Bauml JM, Alley EW, Evans TL, et al. Pretreatment neutrophil-to-lymphocyte ratio as a marker of outcomes in nivolumab-treated patients with advanced non-small-cell lung cancer. Lung Cancer. 2017;106:1–7. Epub 2017/03/14. doi: 10.1016/j.lungcan.2017.01.013. PubMed PMID: 28285682.

17. Diem S, Schmid S, Krapf M, Flatz L, Born D, Jochum W, et al. Neutrophil-to-Lymphocyte ratio (NLR) and Platelet-to-Lymphocyte ratio (PLR) as prognostic markers in patients with non-small cell lung cancer (NSCLC) treated with nivolumab. Lung Cancer. 2017;111:176–81. Epub 2017/08/26. doi: 10.1016/j.lungcan.2017.07.024. PubMed PMID: 28838390.

18. Zahorec R. Neutrophil-to-lymphocyte ratio, past, present and future perspectives. Bratisl Lek Listy. 2021;122(7):474–88. Epub 2021/06/24. doi: 10.4149/bll_2021_078. PubMed PMID: 34161115.

19. Kas B, Talbot H, Ferrara R, Richard C, Lamarque JP, Pitre-Champagnat S, et al. Clarification of Definitions of Hyperprogressive Disease During Immunotherapy for Non-Small Cell Lung Cancer. JAMA Oncol. 2020;6(7):1039–46. Epub 2020/06/12. doi: 10.1001/jamaoncol.2020.1634. PubMed PMID: 32525513; PubMed Central PMCID: PMCPMC7290708 advisory role, or lectures for GE Healthcare, Heartflow, Inc, and Sanofi SA and receiving honoraria from Qualia Systems and WeDiagnostiX. Dr Ferrara reported receiving personal fees Merck Sharp & Dohme. Dr Planchard reported performing consulting, advisory role, or lectures for AstraZeneca plc, Bristol-Myers Squibb, Boehringer Ingelheim, Celgene Corporation, Daiichi Sankyo Company, Limited, Eli Lilly and Company, Merck & Co, Novartis International AG, Pfizer, Inc, prIME Oncology, PeerCME, and Roche Diagnostics; honoraria from AstraZeneca plc, Bristol-Myers Squibb, Boehringer Ingelheim, Celgene Corporation, Eli Lilly and Company, Merck & Co, Novartis International AG, Pfizer, Inc, prIME Oncology, PeerCME, and Roche Diagnostics; clinical trials research for AstraZeneca plc, Bristol-Myers Squibb, Boehringer Ingelheim, Eli Lilly and Company, Merck & Co, Novartis International AG, Pfizer, Inc, Roche Diagnositcs, MedImmune, LLC, Sanofi-Aventis, Taiho Pharmaceutical Co, Ltd, NovoCure Limited, and Daiichi Sankyo Company, Limited; and travel, accommodations, and/or expenses from AstraZeneca plc, Roche Diagnositcs, Novartis International AG, prIME Oncology, and Pfizer, Inc. Dr Besse reported performing sponsored research at Gustave Roussy Cancer Center, AbbVie, Inc, Amgen, Inc, AstraZeneca plc, Biogen, Inc, Blueprint Medicines Corporation, Bristol-Myers Squibb, Celgene Corporatoin, Eli Lilly and Company, GlaxoSmithKline plc, Ignyta, Inc, Ipsen, Merck KGaA, Merck Sharp & Dohme, Nektar Therapeutics, Onxeo, Pfizer, Inc, PharmaMar, Sanofi SA, Spectrum Pharmaceuticals, Takeda Pharmaceutical Company Limited, and Tiziana Pharma, Ltd. Dr Mezquita reported performing consulting or an advisory role for Roche Diagnostics, Roche, and Takeda Pharmaceutical Company Limited; lectures and educational activities for Bristol-Myers Squibb and Tecnofarma SA; and receiving travel, accommodations, and/or expenses from Roche Diagnostics and Bristol-Myers Squibb. Dr Caramella reported receiving personal fees Merck & Co, Bristol-Myers Squibb, and Pfizer, Inc. No other disclosures were reported.

20. Wolchok JD, Hoos A, O’Day S, Weber JS, Hamid O, Lebbé C, et al. Guidelines for the evaluation of immune therapy activity in solid tumors: immune-related response criteria. Clin Cancer Res. 2009;15(23):7412–20. Epub 2009/11/26. doi: 10.1158/1078-0432.Ccr-09-1624. PubMed PMID: 19934295.

21. Kim CG, Kim KH, Pyo KH, Xin CF, Hong MH, Ahn BC, et al. Hyperprogressive disease during PD-1/PD-L1 blockade in patients with non-small-cell lung cancer. Ann Oncol. 2019;30(7):1104–13. Epub 2019/04/13. doi: 10.1093/annonc/mdz123. PubMed PMID: 30977778.

22. Rocha P, Ramal D, Ripoll E, Moliner L, Corbera A, Hardy-Werbin M, et al. Comparison of Different Methods for Defining Hyperprogressive Disease in NSCLC. JTO Clin Res Rep. 2021;2(1):100115. Epub 2021/10/01. doi: 10.1016/j.jtocrr.2020.100115. PubMed PMID: 34589976; PubMed Central PMCID: PMCPMC8474364.

23. Matsuo N, Azuma K, Kojima T, Ishii H, Tokito T, Yamada K, et al. Comparative incidence of immune-related adverse events and hyperprogressive disease in patients with non-small cell lung cancer receiving immune checkpoint inhibitors with and without chemotherapy. Invest New Drugs. 2021;39(4):1150–8. Epub 2021/01/24. doi: 10.1007/s10637-021-01069-7. PubMed PMID: 33483882.

24. Wang X, Mi X, Li T, Li C. Hyperprogressive disease after immune checkpoint inhibitor therapy in a patient with non-small cell lung cancer who harbors a TGFBR2 mutation: A case report. Exp Ther Med. 2023;25(5):228. Epub 2023/04/28. doi: 10.3892/etm.2023.11927. PubMed PMID: 37114179; PubMed Central PMCID: PMCPMC10126802.

25. Zheng Z, Wu K, Yao Z, Mu X, Wu H, Zhao W, et al. Hyperprogressive disease in patients with advanced renal cell carcinoma: a new pattern of post-treatment cancer behavior. Immunol Res. 2020;68(4):204–12. Epub 2020/07/12. doi: 10.1007/s12026-020-09138-4. PubMed PMID: 32651873.

26. Chen Y, Hu J, Bu F, Zhang H, Fei K, Zhang P. Clinical characteristics of hyperprogressive disease in NSCLC after treatment with immune checkpoint inhibitor: a systematic review and meta-analysis. BMC Cancer. 2020;20(1):707. Epub 2020/07/31. doi: 10.1186/s12885-020-07206-4. PubMed PMID: 32727409; PubMed Central PMCID: PMCPMC7392646.

27. Tumeh PC, Hellmann MD, Hamid O, Tsai KK, Loo KL, Gubens MA, et al. Liver Metastasis and Treatment Outcome with Anti-PD-1 Monoclonal Antibody in Patients with Melanoma and NSCLC. Cancer Immunol Res. 2017;5(5):417–24. Epub 2017/04/16. doi: 10.1158/2326-6066.Cir-16-0325. PubMed PMID: 28411193; PubMed Central PMCID: PMCPMC5749922.

28. Castello A, Rossi S, Mazziotti E, Toschi L, Lopci E. Hyperprogressive Disease in Patients with Non-Small Cell Lung Cancer Treated with Checkpoint Inhibitors: The Role of (18)F-FDG PET/CT. J Nucl Med. 2020;61(6):821–6. Epub 2019/12/22. doi: 10.2967/jnumed.119.237768. PubMed PMID: 31862803.

29. Choi YJ, Kim T, Kim EY, Lee SH, Kwon DS, Chang YS. Prediction model for hyperprogressive disease in non-small cell lung cancer treated with immune checkpoint inhibitors. Thorac Cancer. 2020;11(10):2793–803. Epub 2020/08/12. doi: 10.1111/1759-7714.13594. PubMed PMID: 32779394; PubMed Central PMCID: PMCPMC7529559.

30. Refae S, Gal J, Brest P, Giacchero D, Borchiellini D, Ebran N, et al. Hyperprogression under Immune Checkpoint Inhibitor: a potential role for germinal immunogenetics. Sci Rep. 2020;10(1):3565. Epub 2020/02/29. doi: 10.1038/s41598-020-60437-0. PubMed PMID: 32107407; PubMed Central PMCID: PMCPMC7046673 Fréderic Peyrade is a member of an advisory board at M.S.D. and Merck. Delphine Borchiellini is a member of an advisory board at M.S.D., Pfizer, Astra-Zeneca, Roche, B.M.S. Joel Guigay is a member of an advisory board at Merck. The remaining authors declare no competing interests.

31. Vaidya P, Bera K, Patil PD, Gupta A, Jain P, Alilou M, et al. Novel, non-invasive imaging approach to identify patients with advanced non-small cell lung cancer at risk of hyperprogressive disease with immune checkpoint blockade. J Immunother Cancer. 2020;8(2). Epub 2020/10/15. doi: 10.1136/jitc-2020-001343. PubMed PMID: 33051342; PubMed Central PMCID: PMCPMC7555103.

32. Kim Y, Kim CH, Lee HY, Lee SH, Kim HS, Lee S, et al. Comprehensive Clinical and Genetic Characterization of Hyperprogression Based on Volumetry in Advanced Non-Small Cell Lung Cancer Treated With Immune Checkpoint Inhibitor. J Thorac Oncol. 2019;14(9):1608–18. Epub 2019/06/14. doi: 10.1016/j.jtho.2019.05.033. PubMed PMID: 31195179.

33. Matos I, Garralda E. Clarification of Definitions of Hyperprogressive Disease During Immunotherapy. JAMA Oncol. 2021;7(1):136–7. Epub 2020/11/20. doi: 10.1001/jamaoncol.2020.5582. PubMed PMID: 33211082.

34. Matos I, Martin-Liberal J, García-Ruiz A, Hierro C, Ochoa de Olza M, Viaplana C, et al. Capturing Hyperprogressive Disease with Immune-Checkpoint Inhibitors Using RECIST 1.1 Criteria. Clin Cancer Res. 2020;26(8):1846–55. Epub 2019/11/24. doi: 10.1158/1078-0432.Ccr-19-2226. PubMed PMID: 31757877.

35. Ferrara R, Mezquita L, Texier M, Lahmar J, Audigier-Valette C, Tessonnier L, et al. Comparison of Fast-Progression, Hyperprogressive Disease, and Early Deaths in Advanced Non-Small-Cell Lung Cancer Treated With PD-1/PD-L1 Inhibitors or Chemotherapy. JCO Precis Oncol. 2020;4:829–40. Epub 2020/11/01. doi: 10.1200/po.20.00021. PubMed PMID: 35050757.

36. Nishino M, Ramaiya NH, Chambers ES, Adeni AE, Hatabu H, Jänne PA, et al. Immune-related response assessment during PD-1 inhibitor therapy in advanced non-small-cell lung cancer patients. J Immunother Cancer. 2016;4:84. Epub 2016/12/27. doi: 10.1186/s40425-016-0193-2. PubMed PMID: 28018599; PubMed Central PMCID: PMCPMC5168591.

37. Galldiks N, Langen KJ, Pope WB. From the clinician’s point of view - What is the status quo of positron emission tomography in patients with brain tumors? Neuro Oncol. 2015;17(11):1434–44. Epub 2015/07/02. doi: 10.1093/neuonc/nov118. PubMed PMID: 26130743; PubMed Central PMCID: PMCPMC4648307.

38. Chae YK, Wang S, Nimeiri H, Kalyan A, Giles FJ. Pseudoprogression in microsatellite instability-high colorectal cancer during treatment with combination T cell mediated immunotherapy: a case report and literature review. Oncotarget. 2017;8(34):57889–97. Epub 2017/09/17. doi: 10.18632/oncotarget.18361. PubMed PMID: 28915720; PubMed Central PMCID: PMCPMC5593692.

39. Lo Russo G, Moro M, Sommariva M, Cancila V, Boeri M, Centonze G, et al. Antibody-Fc/FcR Interaction on Macrophages as a Mechanism for Hyperprogressive Disease in Non-small Cell Lung Cancer Subsequent to PD-1/PD-L1 Blockade. Clin Cancer Res. 2019;25(3):989–99. Epub 2018/09/13. doi: 10.1158/1078-0432.Ccr-18-1390. PubMed PMID: 30206165.

40. Li G, Choi JE, Kryczek I, Sun Y, Liao P, Li S, et al. Intersection of immune and oncometabolic pathways drives cancer hyperprogression during immunotherapy. Cancer Cell. 2023;41(2):304–22.e7. Epub 2023/01/14. doi: 10.1016/j.ccell.2022.12.008. PubMed PMID: 36638784.

41. Kamada T, Togashi Y, Tay C, Ha D, Sasaki A, Nakamura Y, et al. PD-1(+) regulatory T cells amplified by PD-1 blockade promote hyperprogression of cancer. Proc Natl Acad Sci U S A. 2019;116(20):9999–10008. Epub 2019/04/28. doi: 10.1073/pnas.1822001116. PubMed PMID: 31028147; PubMed Central PMCID: PMCPMC6525547.

42. Camelliti S, Le Noci V, Bianchi F, Moscheni C, Arnaboldi F, Gagliano N, et al. Mechanisms of hyperprogressive disease after immune checkpoint inhibitor therapy: what we (don’t) know. J Exp Clin Cancer Res. 2020;39(1):236. Epub 2020/11/11. doi: 10.1186/s13046-020-01721-9. PubMed PMID: 33168050; PubMed Central PMCID: PMCPMC7650183.

43. Solaymani-Mohammadi S, Lakhdari O, Minev I, Shenouda S, Frey BF, Billeskov R, et al. Lack of the programmed death-1 receptor renders host susceptible to enteric microbial infection through impairing the production of the mucosal natural killer cell effector molecules. J Leukoc Biol. 2016;99(3):475–82. Epub 2015/10/16. doi: 10.1189/jlb.4A0115-003RR. PubMed PMID: 26467188; PubMed Central PMCID: PMCPMC6608046.

44. Lamichhane P, Karyampudi L, Shreeder B, Krempski J, Bahr D, Daum J, et al. IL10 Release upon PD-1 Blockade Sustains Immunosuppression in Ovarian Cancer. Cancer Res. 2017;77(23):6667–78. Epub 2017/10/11. doi: 10.1158/0008-5472.Can-17-0740. PubMed PMID: 28993412; PubMed Central PMCID: PMCPMC5712245.

