## supplementary figures for "Redefining Clinical Hyperprogression: the Incidence, Clinical Implications, and Risk Factors of Hyperprogression in Non-Small Cell Lung Cancer Treated with Immunotherapy"

**Supplementary Figure 1.**

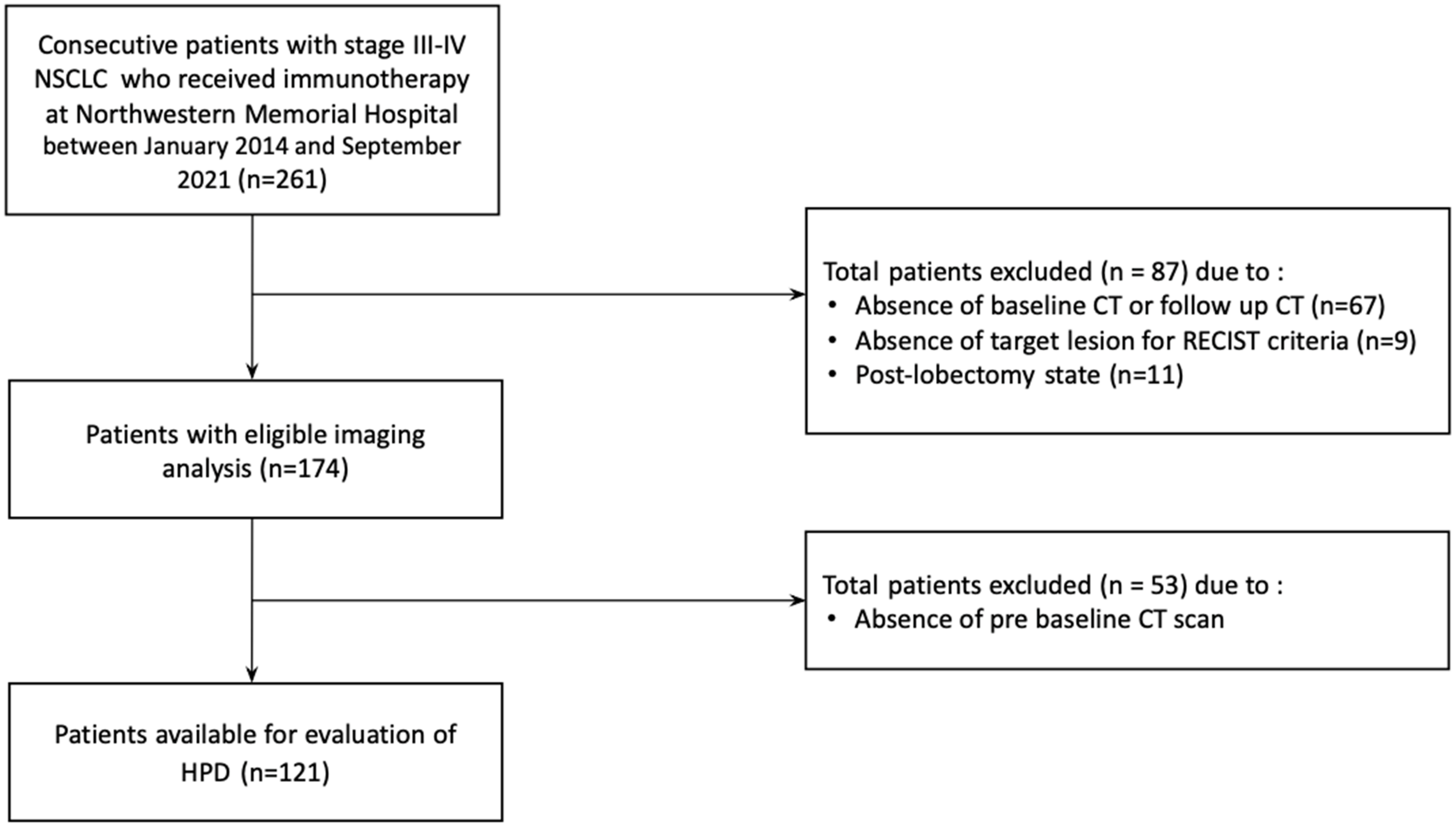

**Supplementary Figure 2.**

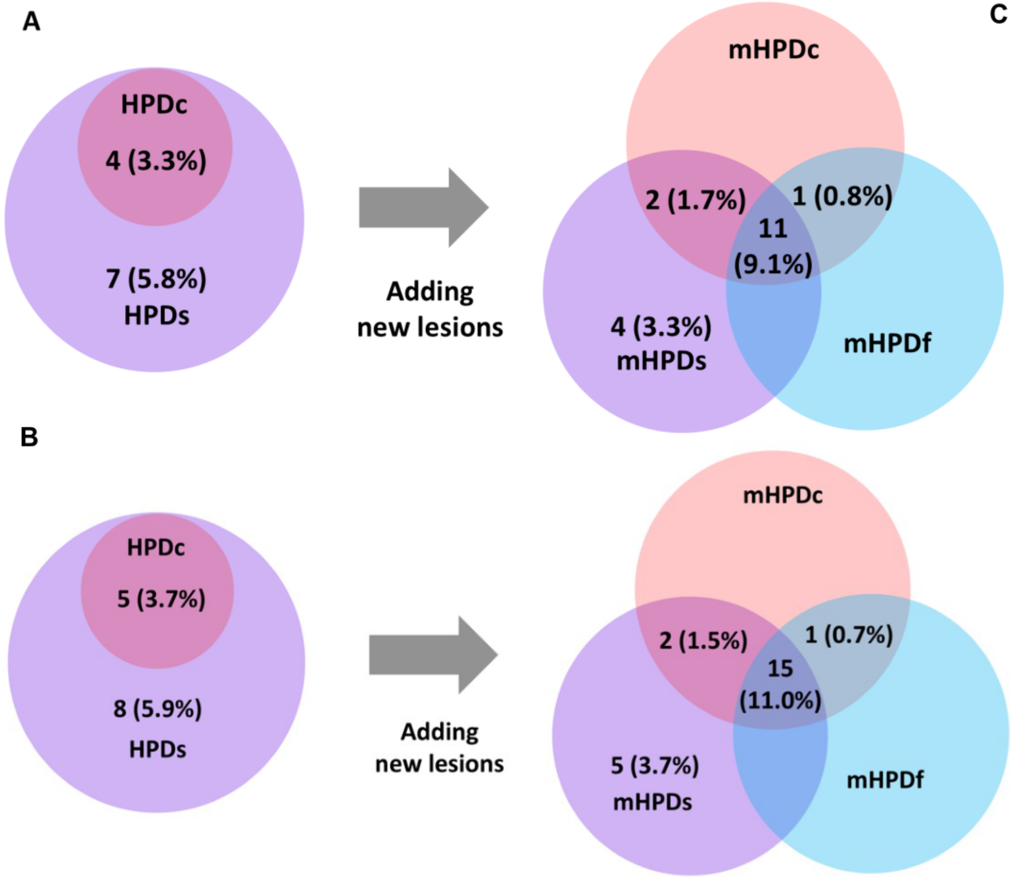


C

| Definitions of HPD | Using parameters | Cases of patient (n=121) | Cases of immunotherapy (n=136) |
|---|---|---|---|
| HPDc | RECIST progression, TGR-exp/TGR-ref >2 | 4 (3.3) | 5 (2.9) |
| mHPDc | HPDc + new lesions | 14 (11.6) | 18 (13.2) |
| HPDs | TGK-exp/TGK-ref>2 | 11 (9.1) | 14 (10.3) |
| mHPDs | HPDs + new lesions | 17 (14.0) | 22 (16.2) |
| HPDf | RECIST progression, TGR-exp/TGR-ref>50% | 0 (0.0) | 0 (0.0) |
| mHPDf | HPDf + new lesions | 12 (9.9) | 16 (11.8) |

**Supplementary Figure 3.**

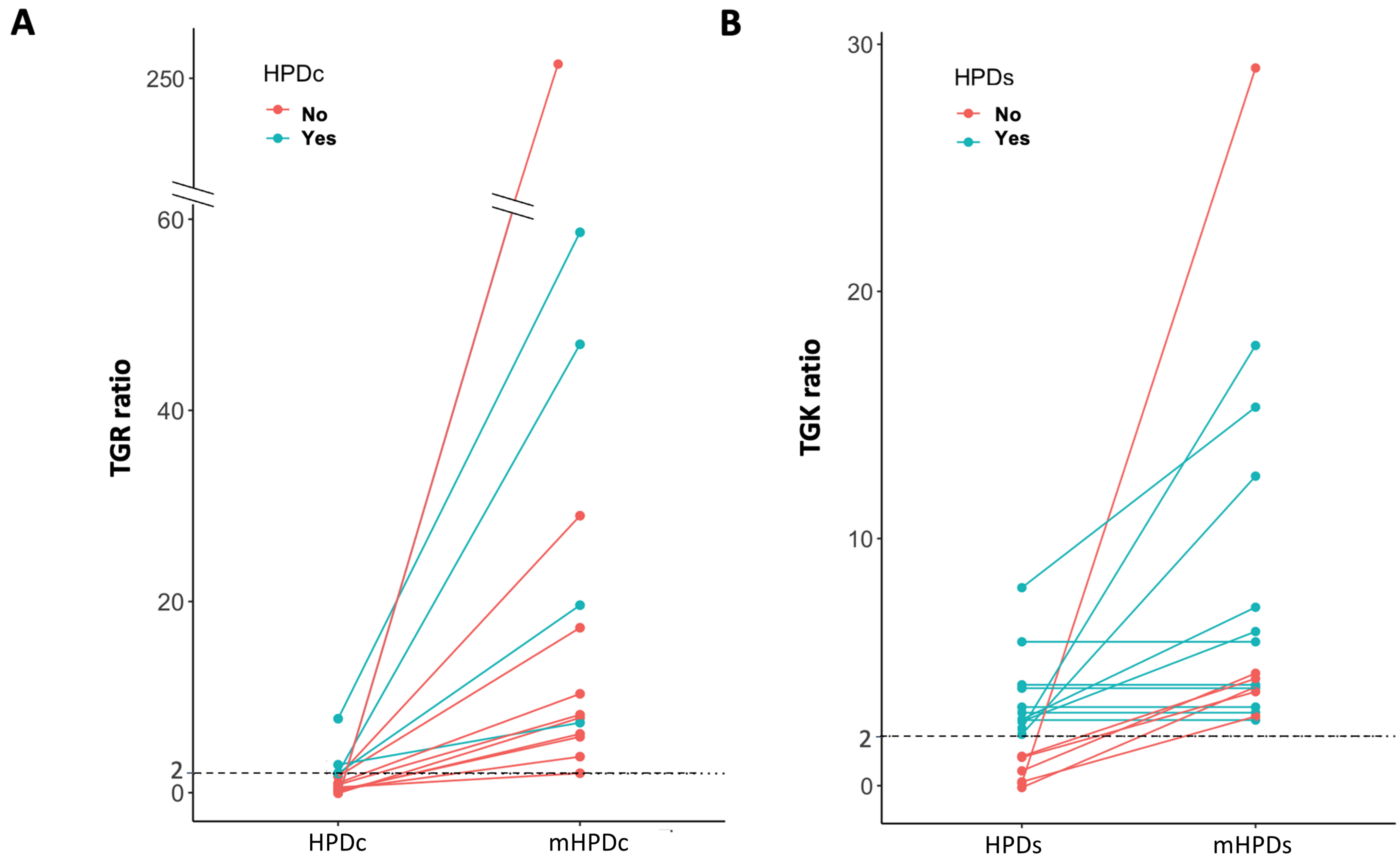

**Supplementary Figure 4.**

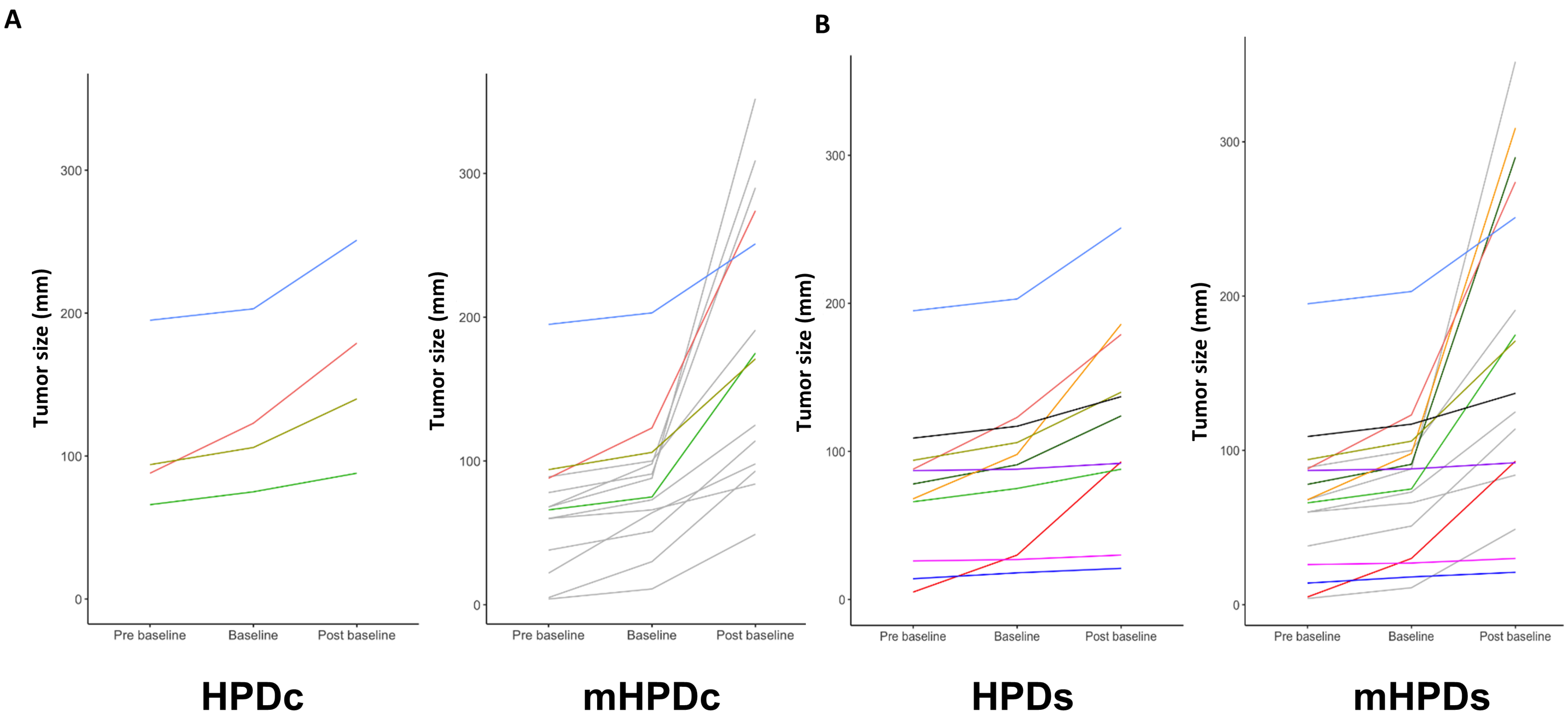

**Supplementary Figure 5.**

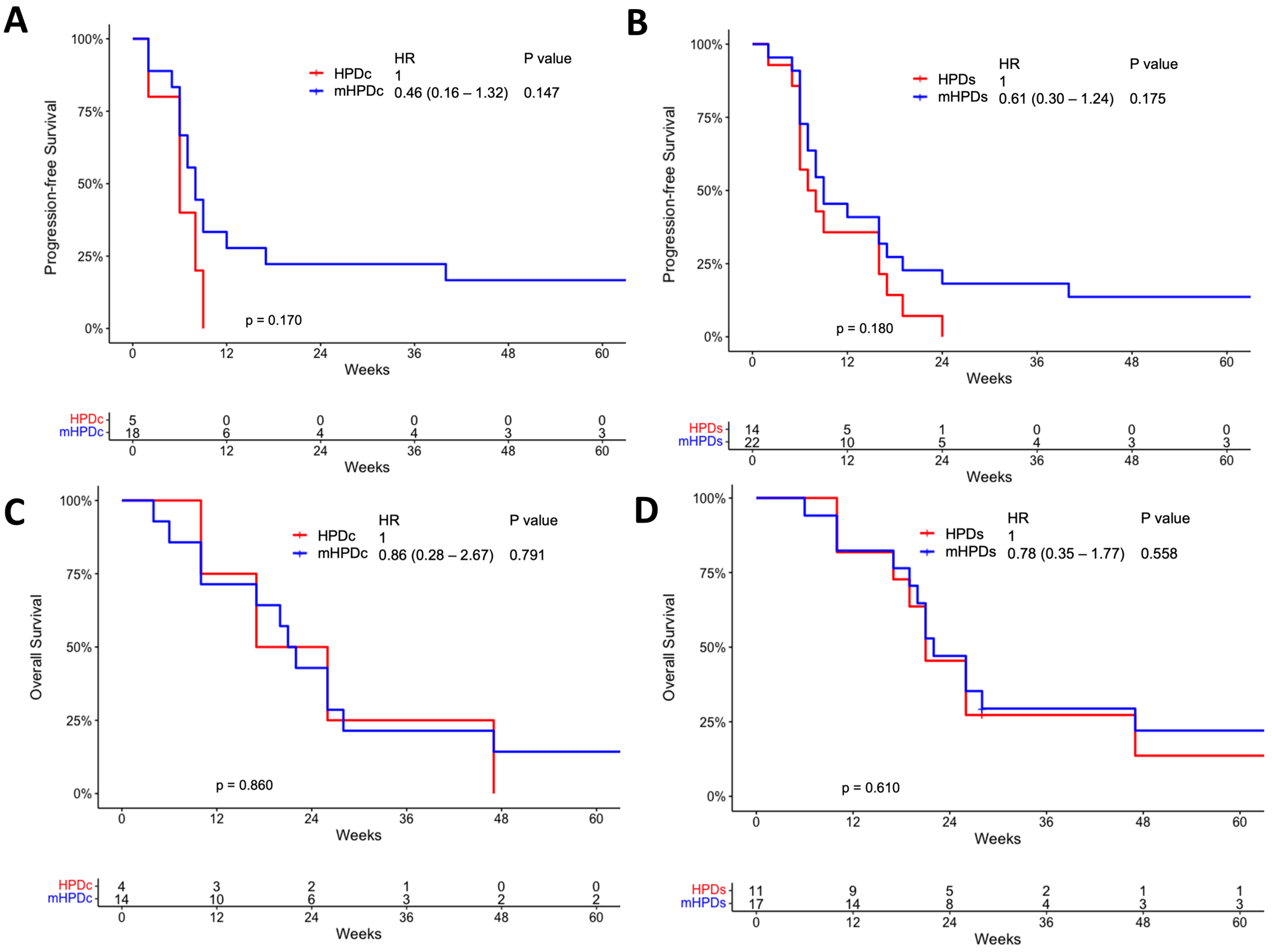
